# A prospective, comparative cohort analysis of influenza antibody waning in Michigan and Hong Kong during periods of low influenza circulation

**DOI:** 10.1101/2025.05.26.25328346

**Authors:** Yanyupei Yang, Matthew Smith, Faith Ho, Rachel Truscon, Nancy H. L. Leung, Lisa Touyon, Amy Callear, Elie-Tino Godonou, Christopher N. Blair, Arnold S. Monto, Adam S. Lauring, Benjamin J. Cowling, Sook-San Wong, Emily T. Martin

**Author notes:** Co-corresponding authors: Yangyupei Yang, address: 1415 Washington Heights, Ann Arbor, MI, USA, 48109. Emily T. Martin, address: 1415 Washington Heights, Ann Arbor, MI, USA, 48109. Joint senior authors (Sook-San Wong, Emily T. Martin).

## Abstract

**Background:** Reduced influenza transmission during the COVID-19 pandemic prompted concern about waning of population immunity that could lead to subsequent surges in circulation. We compared longitudinal influenza antibody titers in Michigan and Hong Kong, two regions with reduced influenza transmission during the COVID-19 pandemic.

**Methods:** In two prospective cohort studies (HIVE, Michigan; EPI-HK, Hong Kong), we analyzed longitudinal serum samples collected from 2020 through 2023 from participants without influenza infection or vaccination. Sera were tested using hemagglutination inhibition assays (HAI) and microneutralization (MN) against vaccine strains. Two-year fold changes in titers were estimated using multivariable power-law models adjusted for age and baseline titer.

**Results:** We analyzed 173 sera from 57 HIVE participants and 259 sera from 60 EPI-HK participants. Initial HAI geometric mean titers ranged from 12.3–123.4 in HIVE and 6.3–40.9 in EPI-HK. Among those with >1:40 baseline titers in HIVE, fold-change estimates indicated waning for H1N1 strains, including A/Brisbane (0.70 [95% CI: 0.56,0.86]), A/Idaho (0.40 [0.32,0.50]), and A/Michigan (0.43 [0.34,0.52]), while H3N2 and influenza B showed minimal or no waning. In EPI-HK, fold-change estimates demonstrated heterogeneous patterns by strain for H1N1, and waning for selected H3N2 strains, including A/Cambodia (0.44 [0.29,0.65]) and A/Singapore (0.48 [0.33,0.69]). MN results were consistent with HAI findings.

**Conclusion:** Influenza antibody responses during the pandemic showed heterogeneous, strain-specific changes rather than uniform decline. These findings suggest that population immunity was maintained for many strains despite reduced viral circulation, with waning occurring in particular strains and varying by population.

**Summary:** Influenza antibody responses during the pandemic showed heterogeneous, strain-specific changes rather than uniform decline in Hong Kong and Michigan.

Adam S. Lauring holds the position of Deputy Editor for JID and has not peer reviewed or made any editorial decisions for this paper.

## Background

The COVID-19 pandemic and implementation of mitigation strategies changed the global circulation of respiratory viruses, including seasonal influenza. Interventions such as international travel restrictions, school closures, masking, and social distancing led to low influenza activity between 2020 and 2022, with some regions reporting absence of influenza transmission^1–3^. While these interventions effectively reduce transmission, concerns remained regarding longer-term impacts on population immunity to respiratory viruses. As a consequence of influenza’s seasonality, waning of antibody levels has long been a topic of interest, particularly in informing vaccine timing and frequency. However, it has been challenging to isolate the role of natural infection in antibody waning, given frequent infections, even among vaccinated individuals. The COVID-19 pandemic provided a unique opportunity to study antibody waning in near absence of influenza virus exposure^4–7^.

In US, influenza activity demonstrated a clear seasonal pattern prior to the COVID-19 pandemic, typically beginning in November and continuing through March each year^8^. In response to rising COVID-19 cases, Michigan implemented a series of early and aggressive mitigation measures in March 2020, including statewide K–12 school closures beginning March 12, followed by the “Stay Home, Stay Safe” executive order issued on March 23^9^. These interventions, which remained in place until June 1, 2020, were associated with substantial reductions in SARS-CoV-2 transmission and coincided with marked disruption of 2019–2020 and 2020–2021 influenza seasons. Following implementation of these measures, including school closures, widespread masking, social distancing, and heightened public awareness, no influenza cases were detected in the cohort from March 2020 through October 2021. Influenza transmission re-emerged during 2021–2022 season, dominated by H3N2 strains.

In Hong Kong, universal masking was mandated from mid 2020 until early 2023. Social distancing measures were introduced in early 2020, including restrictions on public gatherings, closures of high-risk venues, and dining limitations. Schools were closed in late January 2020 and shifted to remote learning, with intermittent reopenings followed by reclosures during subsequent waves. Quarantine for inbound travelers began in February 2020 and remained in effect until September 2022^10^. These prolonged public health measures were associated with very low levels of influenza activity, and no community-detected influenza cases were reported through surveillance systems between March 2020 and January 2023. Influenza began to spread again in Hong Kong in early 2023 after the relaxation of a mask mandate, with influenza circulation initially predominated by H1N1 viruses^11^, followed by H3N2 strains ^12^.

To address these questions, we evaluated longitudinal antibody responses in two prospective cohort studies: the Household Influenza Vaccine Evaluation (HIVE) cohort in Michigan, United States, and the Evaluating Population Immunity in Hong Kong (EPI-HK) cohort. These cohorts represent two distinct epidemiological and public health contexts: Michigan experienced moderate influenza vaccination uptake and a relatively early relaxation of COVID-19 mitigation measures^9^, while Hong Kong maintained stringent control policies for longer, including prolonged mask mandates, quarantine protocols, and reduced social mixing^10^. By comparing selected participants who did not receive influenza vaccination across two distinct epidemiological and cultural settings, we aimed to gain unique insights into durability of influenza immunity during this unique period.

## Methods

### Participant Enrollment and Data Collection

#### HIVE Cohort

The Household Influenza Vaccine Evaluation (HIVE) study is a prospective cohort of households with children, conducted in Ann Arbor, Michigan and surrounding areas. Recruitment into cohort occurred annually, with households committing to one year of participation at a time while being encouraged to participate for multiple seasons^8^. For this analysis, Participants were selected using an age-balanced sampling approach restricted to unvaccinated individuals during July 2020 December 2022 (supplementary methods). Specifically, participants were stratified into age groups (0–17, 18–29, 30–44, 45–54, and 55–64 years), with a target of approximately 20 participants per stratum when available; if fewer than 20 eligible participants were available in a given age group, all unvaccinated individuals in that stratum were included (Table S1).

Each year, participants completed an enrollment survey and were prospectively followed for acute respiratory illness (ARI) symptoms. Household members who reported at least two ARI symptoms were asked to provide combined nasal and throat swab specimens, which were tested for influenza using reverse transcription polymerase chain reaction (RT-PCR)^8^. Since 2011, blood samples have been collected twice annually from participants. Pre-season serum specimens were collected in Fall and post-season serum specimens were collected in Summer.

#### EPI-HK Cohort

The Evaluating Population Immunity in Hong Kong (EPI-HK) study is a longitudinal community-based cohort study involving intensive follow-up of approximately 2000 individuals of all ages enrolled fromgeneral community in Hong Kong^13^. Participants are followed up with blood draws every 6 months, at which time information on health status, vaccination history, and hospitalization are also collected. For this analysis, participants were similarly selected across age groups as in HIVE cohort (Supplementary methods). Ten participants were randomly selected within each age stratum (0–17, 18–29, 30–44, 45– 54, 55–64, 65–74 years), and an additional up to 30 participants per age group were included for sensitivity analyses (Table S1). Serum specimens collected among selected participants between July 2020 and December 2022 were included.

### Laboratory Methods

All serum specimens were tested for antibody responses against each influenza vaccine strain using hemagglutination inhibition (HAI) assays in each cohort. Antigens representing seasonally relevant vaccine strains were used for testing across both cohorts (Table 1, Table S2).

**A prospective, comparative cohort analysis of influenza antibody waning in Michigan and Hong Kong during periods of low influenza circulation**

**Table 1:** HAI antigen tested in each cohort.

| Influenza Virus Strain | HIVE HAI antigen (year included in seasonal vaccination<sup>1</sup>) | EPI-HK HAI antigen (year included in seasonal vaccination<sup>1</sup>) | HIVE LMN antigen | EPI-HK LMN antigen |
| --- | --- | --- | --- | --- |
| **A(H1N1)** | A/Michigan/45/2015 (2017-2019)<sup>2</sup>,<br>A/Brisbane/2/2018 (2019-2020),<br>A/Idaho/07/2018 (2019-2020)<sup>2</sup> | A/Michigan/45/2015 (2017-2019),<br>A/Brisbane/2/2018 (2019-2020),<br>A/Hawaii/70/2019 (2020-2022)<sup>2</sup>,<br>A/Victoria/2570/2019 (2021-2022), | A/Michigan/45/2015 (2017-2019),<br>A/Idaho/07/2018 (2019-2020)<sup>2</sup> | A/Michigan/45/2015 (2017-2019) |
| **A(H3N2)** | A/Singapore/INFIMH-16-0019/2016 (2018-2019)<sup>2</sup>,<br>A/Kansas/14/2017 (2019-2020)<sup>2</sup> | A/Hong Kong/4801/2014 (2016-2018),<br>A/Singapore/INFIMH-16-0019/2016 (2018-2019),<br>A/Kansas/14/2017 (2019-2020),<br>A/Cambodia/e0826360/2020 (2021-2022) | A/Singapore/INFIMH-16-0019/2016 (2018-2019),<br>A/Kansas/14/2017 (2019-2020), | A/Singapore/INFIMH-16-0019/2016 (2018-2019),<br>A/Kansas/14/2017 (2019-2020), |
| **B/Yamagata** | B/Phuket/3072/2013 (2015-2023) | B/Phuket/3073/2013 (2015-2023) |  |  |
| **B/Victoria** | B/Colorado/06/2017 (2018-2020)<sup>2</sup> | B/Washington/02/2019 (2020-2022) |  |  |

<sup>1</sup>Influenza strain included in the seasonal influenza vaccine formulation for the indicated year.
<sup>2</sup> Cell-culture derived strains

To assess inter-laboratory consistency, a subset of serum samples from EPI-HK cohort (N = 71 samples from 10 participants) was tested in University of Michigan laboratory using HAI assays for three antigens: A/Michigan/45/2015, A/Kansas/14/2017, and A/Singapore/INFIMH-16-0019/2016. In addition, all HIVE serum samples and same subset of EPI-HK serum samples used for cross-laboratory comparison were tested using microneutralization (MN) assays^14^ for same antigens. Serologic testing was performed independently in laboratory at University of Michigan School of Public Health and at University of Hong Kong School of Public Health using standard procedures^15^ (supplementary methods).

### Statistical Methods

Baseline participant and household characteristics were summarized and compared between two cohorts using chi-squared (χ^2^) tests or Fisher’s exact tests for categorical variables. HAI and MN titers were log_2_-transformed, and geometric mean titers (GMT) with 95% confidence intervals (CI) were calculated for each sample collection period in both cohorts. Fold changes in GMT were calculated by comparing titers from the last and first sample collection periods within study period.

To evaluate inter-laboratory agreement, we compared HAI titers measured in University of Michigan and University of Hong Kong laboratories among the subset of EPI-HK samples that were re-tested in both settings. We also assessed agreement between HAI and MN assays. Agreement in antibody titers was assessed using Pearson correlation coefficients for each shared antigen. We additionally evaluated agreement in seroprotection status, defined as HAI titers <1:40 versus ≥1:40, using Cohen’s kappa statistics.

To assess antibody waning, we conducted analyses restricted to participants with baseline HAI titers ≥1:40, a commonly used seroprotection threshold associated with reduced risk of influenza infection^16,17^. For each antigen, log2-transformed HAI or MN titers were modeled as a function of months since baseline using repeated measurements from each participant. We applied three modeling approaches: linear mixed-effects, power law, and natural cubic spline models with a random intercept by participant (supplementary methods). Multivariate models were fitted, adjusting for participant age and baseline titer. Model selection was based on Akaike Information Criterion (AIC) and Bayesian Information Criterion (BIC). Using these models, we estimated antibody trajectories over time and calculated fold changes in predicted titers at two years after baseline compared with baseline levels, adjusting for age and baseline titer. All analyses were conducted using R (version 4.3.1).

## Results

### Characteristics of Households and Participants

A total of 57 participants (173 serum samples) from HIVE cohort and 60 participants (259 serum samples) from EPI-HK cohort were included in analysis (Figure 1, Table 2, Table S1). There was no significant difference in sex distribution between two cohorts (Table 2). Regarding historical influenza vaccination from 2015 to 2019, 74% of HIVE participants and 7% of EPI-HK participants had received vaccine. 32% of HIVE participants had laboratory-confirmed symptomatic influenza virus infections during 2015 to 2019 (Table 2). Infection history of EPI-HK participants was unknown. During 2022-2023 influenza season, 2 (0.2%) HIVE participants were infected and developed symptoms, whereas no infections were reported in EPI-HK cohort. HIVE participants came from larger households than EPI-HK participants; mean numbers of children and adults in HIVE households were 3 (range: 1-6) and 2 (range: 1-6), respectively. For EPI-HK cohort, the averages were 0 (range: 0-3) for children and 3 (range 1-6) for adults.

**Table 2:**
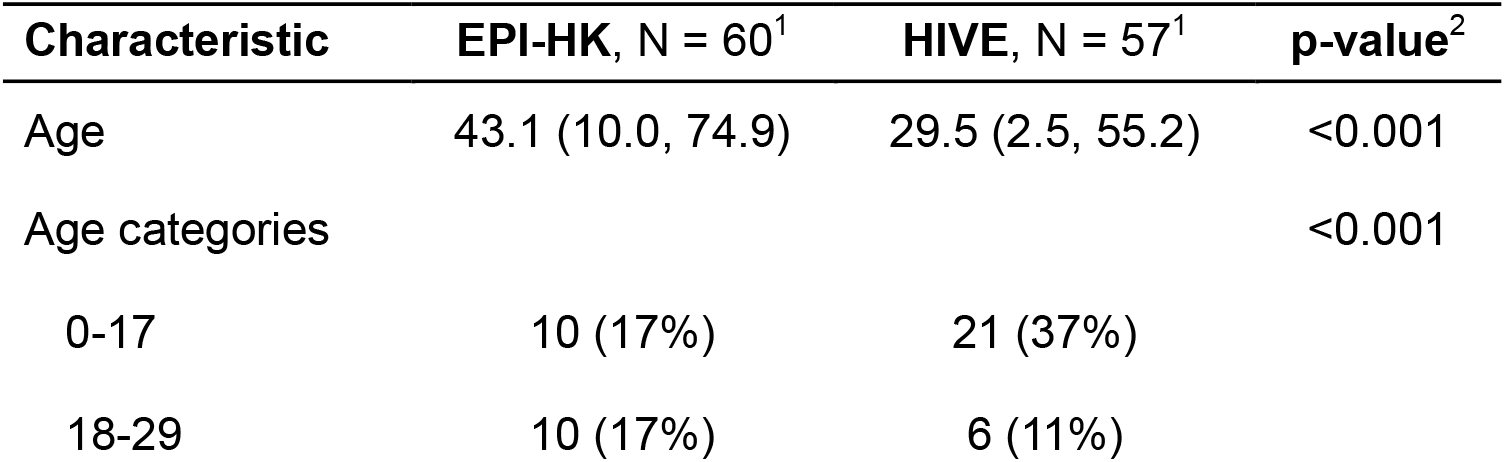
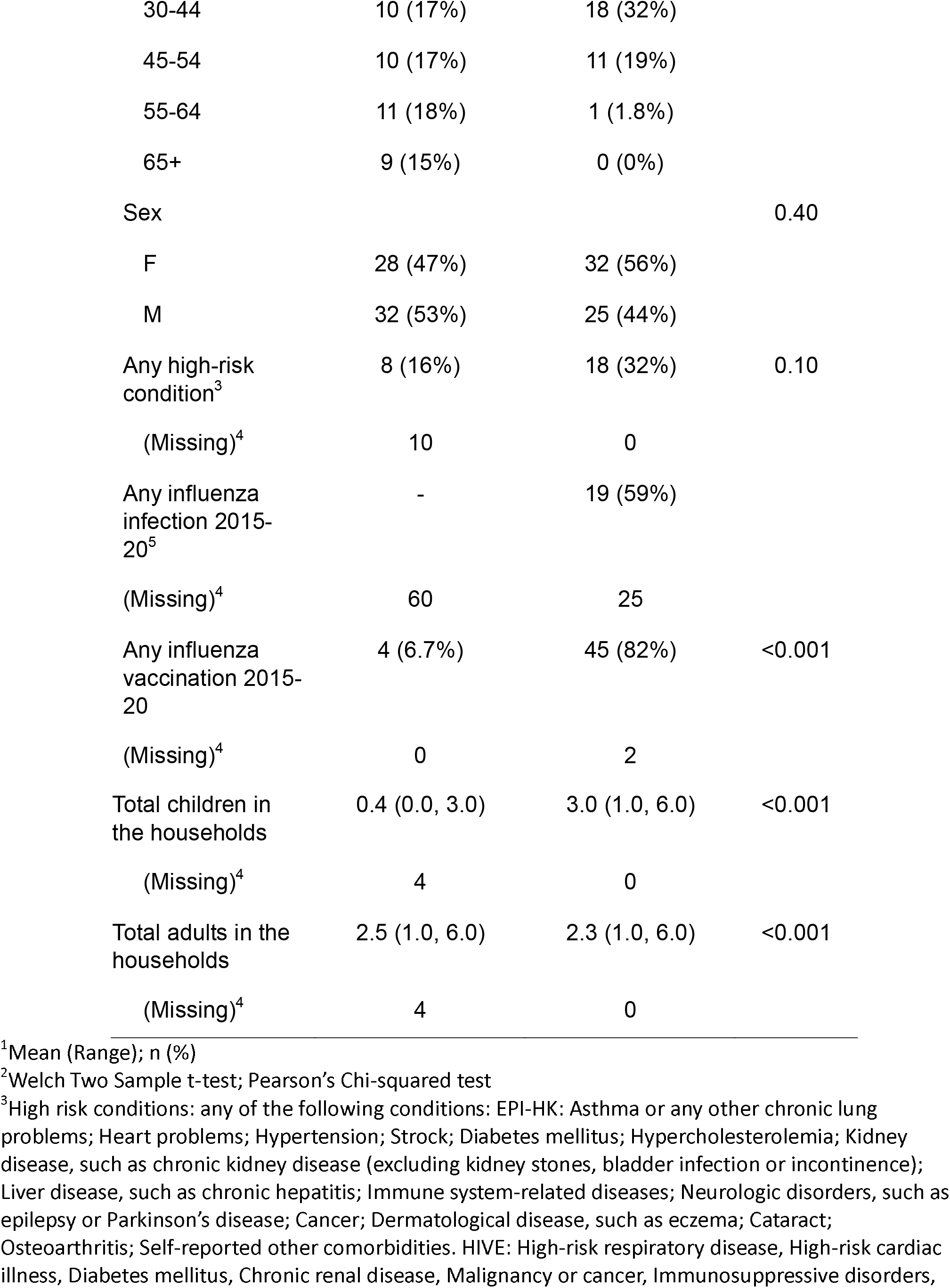
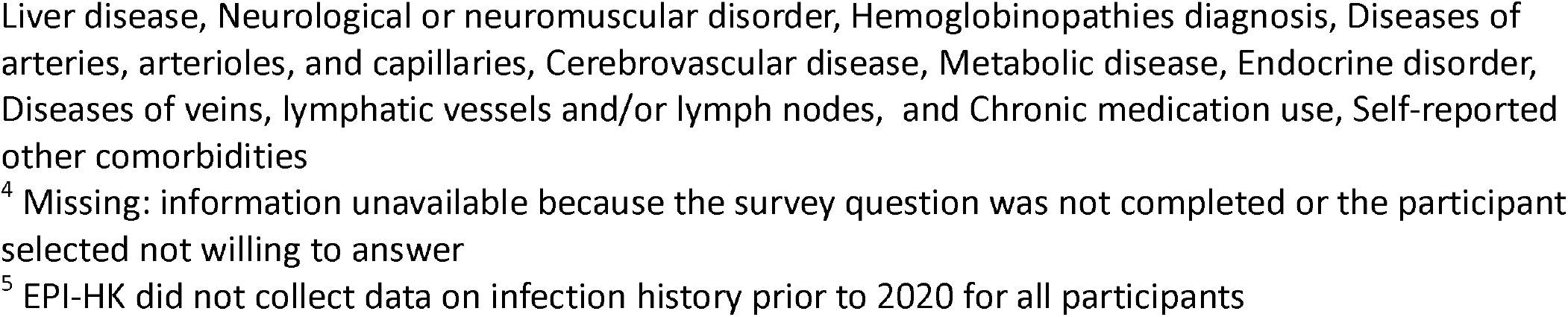
Characteristics of participants.

**Figure 1:**
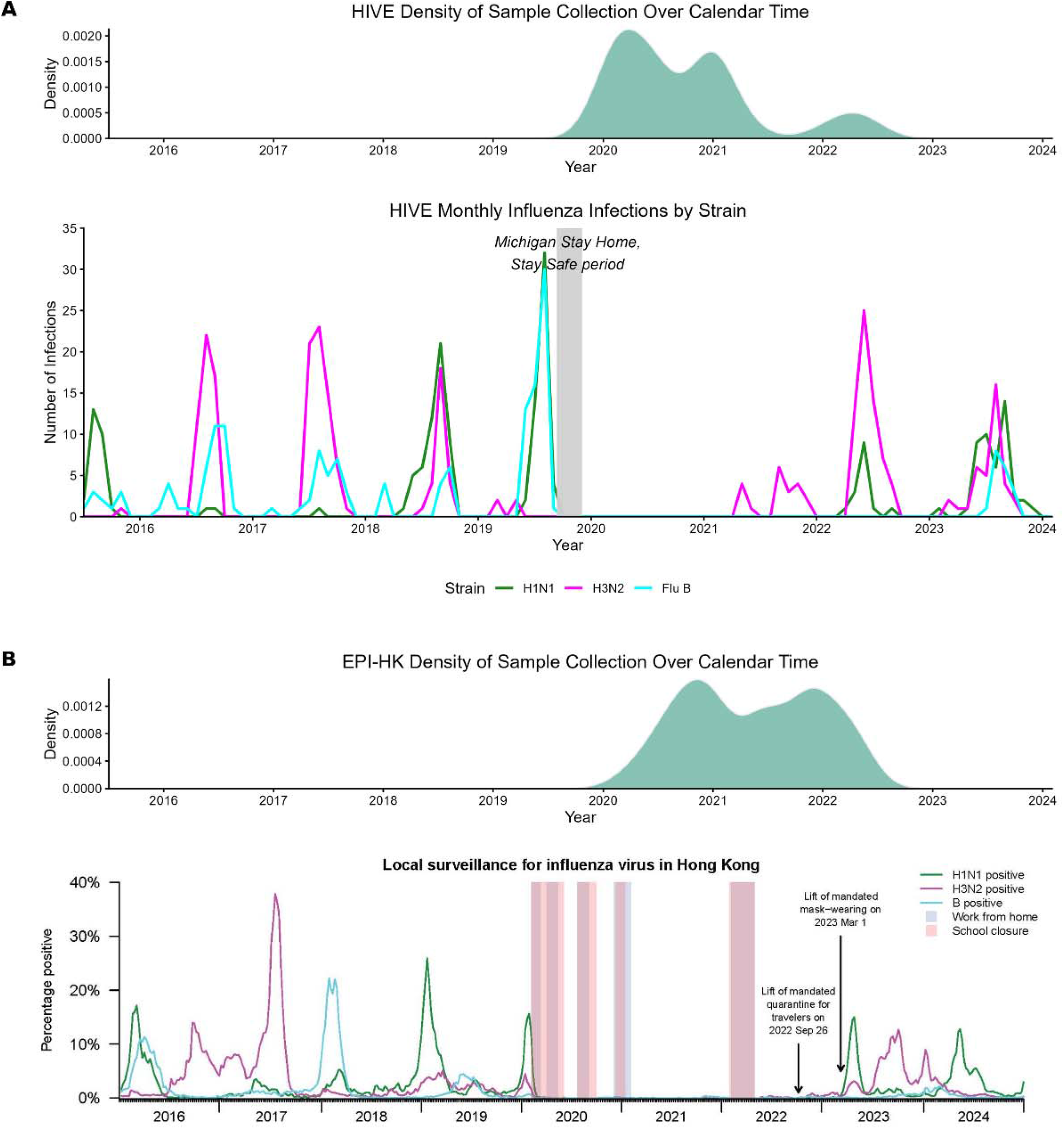
Influenza surveillance trends and serum sample collection density in Michigan and Hong Kong, 2016–2024 A. HIVE cohort (Michigan). The influenza epidemic curve represents laboratory-confirmed influenza infections identified through active surveillance among HIVE participants annually. The serum sample collection density reflects the timing and frequency of serum specimens collected from study participants. B. Hong Kong. The influenza epidemic curve reflects surveillance data from the Hong Kong government laboratory. Percent positivity represents the proportion of respiratory specimens testing positive for influenza among all specimens submitted to the Public Health Laboratory Services Branch of the Centre for Health Protection and the Hospital Authority. These specimens were collected from patients presenting with influenza-like illness in outpatient clinics or during hospitalization. The serum sample collection density reflects cohort-based sampling from EPI-HK participants.

### Baseline HAI and LMN strain-specific antibody levels

At baseline, GMTs measured by HAI were consistently higher in HIVE cohort compared with EPI-HK cohort across all influenza subtypes. For H1N1, baseline GMTs ranged 77.1–106.4 in HIVE (n ≥40 = 44-47) versus 7.2–15.3 in EPI-HK (n ≥40 = 5-18). For H3N2, GMTs ranged 53.8–123.4 in HIVE (n ≥40 = 38-45) and 6.3–17.8 in EPI-HK (n ≥40 = 2-21). And for influenza B, baseline GMTs were 37.1-56.6 in HIVE (n ≥40 = 38-41) and 8.0-11.9 in EPI-HK (n ≥40 = 5-35) (Figure S1, Table S3). Across age groups (<18 vs. ≥18 years), no significant differences in baseline GMTs were observed in either cohort.

Among tested influenza A strains using MN assay, GMTs were consistently higher in HIVE than in EPI-HK across influenza subtypes at baseline (Table S3). For H1N1 viruses, baseline MN GMTs ranged from 399.9 to 656.0 in HIVE for A/Idaho/07/2018 (A/Idaho) and A/Michigan/45/2015 (A/Michigan), compared with 80.0 for A/Michigan in EPI-HK. For H3N2 viruses, baseline MN GMTs ranged from 166.1 to 229.1 in HIVE for A/Singapore/INFIMH-16-0019/2016 (A/Singapore) and A/Kansas/14/2017 (A/Kansas), compared with 62.6 to 68.0 in EPI-HK. Comparison between assay platforms showed strong concordance between HAI and MN titers for H1N1 strains, whereas correlations for H3N2 were low (Figure S3, S4).

Cross-laboratory comparisons demonstrated generally good agreement between HAI assays performed at University of Michigan and University of Hong Kong laboratories. Pearson correlation coefficients were 0.94 for H1N1 A/Michigan, 0.91 for H3N2 A/Singapore, and 0.83 for H3N2 A/Kansas (Figure S2), indicating that both laboratories captured highly similar relative antibody patterns across shared serum specimens. Agreement based on seroprotection threshold (HAI titer ≥1:40) was also substantial for H1N1 A/Michigan (κ = 0.86), moderate for H3N2 A/Singapore (κ = 0.39), and slight for H3N2 A/Kansas (κ = 0.07). Comparison between assay platforms also showed variable agreement between HAI and MN. Concordance was moderate for H1N1 strains (EPI-HK: κ = 0.48; HIVE: κ = 0.51–0.68), whereas agreement for H3N2 was lower (EPI-HK: κ = 0.15–0.46; HIVE: κ = 0.02–0.60) (Figures S3 and S4).

### Antibody waning among participants with >1:40 seroprotection at baseline

In HIVE, 14.3%–28.6% of participants showed increases in HAI titer (A/Kansas and A/Idaho, respectively), compared to 3.3%–23.3% in EPI-HK (A/Singapore and B/Phuket, respectively), likely reflecting additional infections (asymptomatic or too mild to meet case definitions) not captured in active surveillance (Table S3). Among participants included in waning analyses, the proportion with seroprotective HAI titers (≥1:40) at baseline ranged from 67.9%–83.9% across strains in HIVE and from 3.3%–58.3% in EPI-HK, with generally similar proportions remaining seroprotected at final serum collection (Table S4).

Based on AIC and BIC comparisons, power-law models were identified as best-fitting approach for modeling waning rates in all antigens in both cohorts (Table S5-S8). All results presented below are derived from multivariable power-law models adjusted for age and baseline titer and are presented as monthly waning rates and corresponding two-year fold-change estimates.

#### H1N1

Among participants with baseline HAI titers ≥1:40, monthly HAI waning rates for H1N1 strains in HIVE were –0.10 (95% CI: –0.30, 0.11) for A/Brisbane, –0.23 (–0.47, 0.00) for A/Idaho, and –0.22 (–0.40, – 0.03) for A/Michigan (Table S5, Figure S5). Two-year fold-change estimates indicated moderate waning for A/Brisbane (0.70 [0.56,0.86]) and significant waning for A/Idaho (0.40 [0.32,0.50]) and A/Michigan (0.43 [0.34,0.52]), with greatest decline observed for A/Idaho and A/Michigan (Figure 2).

**Figure 2:**
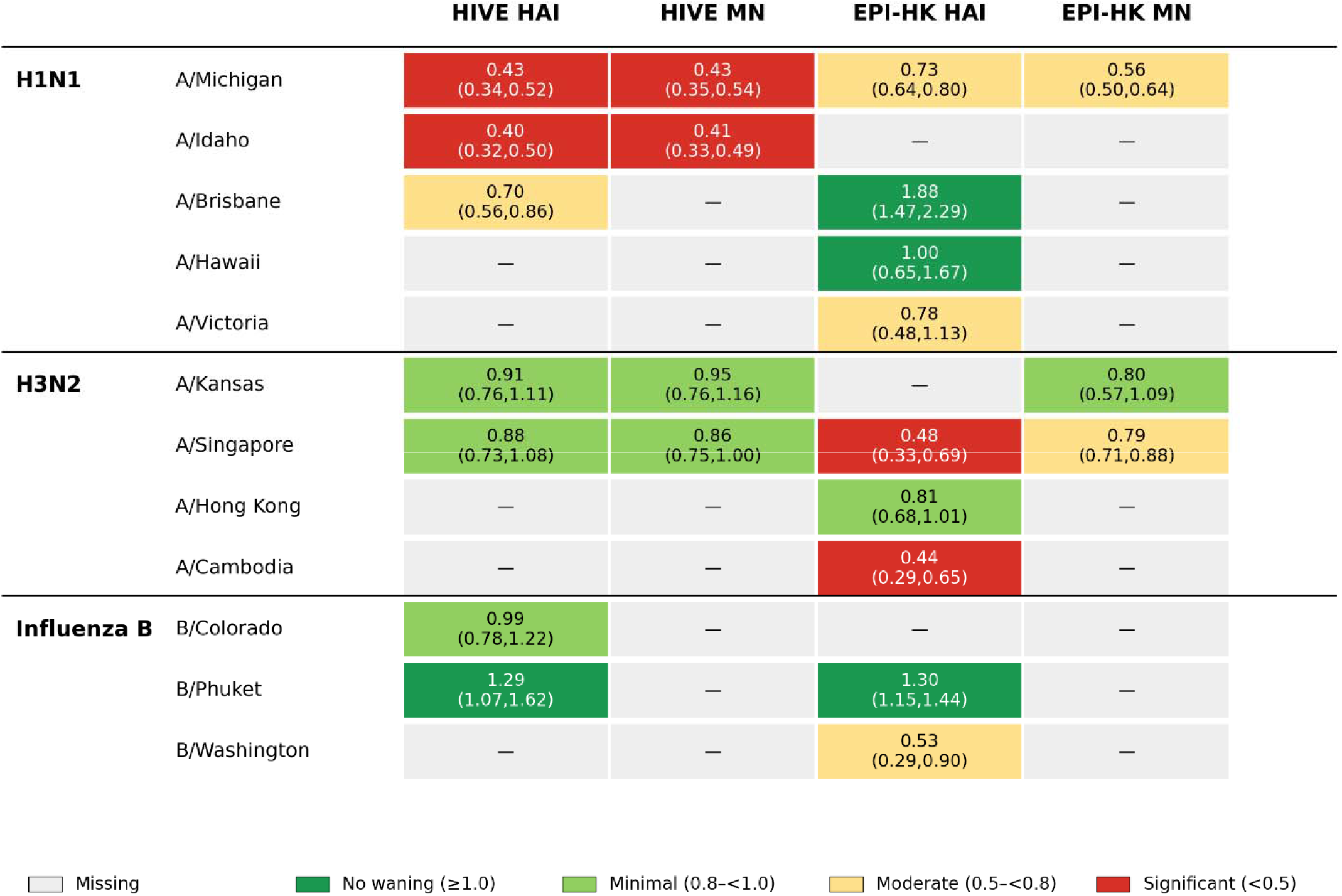
Predicted two-year fold change in influenza antibody titers among participants with baseline seroprotective antibody levels (HAI titer ≥1:40) in the HIVE and EPI-HK cohorts. Values represent predicted fold changes (95% confidence intervals) in hemagglutination inhibition (HAI) and microneutralization (MN) antibody titers two years after baseline, estimated from multivariable power-law mixed-effects models adjusted for age and baseline antibody titer. Analyses were restricted to participants with baseline HAI titers ≥1:40. Strains evaluated included A(H1N1)pdm09 viruses A/Michigan/45/2015, A/Idaho/07/2018, A/Brisbane/02/2018, A/Hawaii/70/2019, and A/Victoria/2570/2019; A(H3N2) viruses A/Kansas/14/2017, A/Singapore/INFIMH-16-0019/2016, A/Hong Kong/4801/2014, and A/Cambodia/e0826360/2020; and influenza B viruses B/Colorado/06/2017, B/Phuket/3073/2013 (Yamagata lineage), and B/Washington/02/2019 (Victoria lineage). Cell colors indicate the magnitude of antibody waning: no waning (fold change ≥1.0), minimal waning (0.8–<1.0), moderate waning (0.5–<0.8), substantial waning (<0.5), and missing that no testing was done (gray).

Using MN assays, similar patterns were observed in HIVE, with estimated monthly declines of –0.17 (– 0.35, 0.01) for A/Michigan and –0.15 (–0.37, 0.08) for A/Idaho (Table S6). Corresponding two-year fold-change estimates indicated significant waning for both A/Michigan (0.43 [0.35,0.54]) and A/Idaho (0.41 [0.33,0.49])(Figure 2).

In EPI-HK, A/Brisbane (0.16 [–0.04,0.35]) and A/Hawaii (0.003 [–0.28,0.23]) showed no evidence of monthly waning, whereas A/Michigan (–0.08 [–0.21,0.04]) and A/Victoria (–0.06 [–0.35,0.22]) showed monthly declines, although confidence intervals included the null (Table S7). Consistent with these results, two-year fold-change estimates indicated no waning for A/Brisbane (1.88 [1.47,2.29]) and A/Hawaii (1.00 [0.65,1.67]), moderate waning for A/Michigan (0.73 [0.64,0.80]), and minimal waning for A/Victoria (0.78 [0.48,1.13]).

MN models in EPI-HK indicated monthly declines for A/H1N1/Michigan (–0.02 [–0.03, –0.003]) (Table S8). Fold-change estimates further supported waning, with a two-year fold change of 0.56 (0.50,0.64), indicating moderate waning.

#### H3N2

Among participants with baseline HAI titers ≥1:40, HAI monthly waning rates in HIVE were –0.02 (– 0.23,0.18) and –0.03 [–0.18, 0.12] for A/Kansas and A/Singapore, showing little change over time. Fold-change estimates were consistent with these findings, with A/Kansas (0.91 [0.76,1.11]) and A/Singapore (0.88 [0.73,1.08]) remaining close to one, indicating minimal waning.

MN models in HIVE showed similar stability, with monthly changes of –0.05 (–0.24, 0.15) for A/Kansas and –0.04 (–0.24,0.15) for A/Singapore, and two-year fold-change estimates were 0.95 (0.76, 1.16) and 0.86 (0.75, 1.00) respectively for A/Kansas and A/Singapore, indicating minimal waning.

In EPI-HK, HAI results indicated monthly declines across H3N2 strains, including A/Cambodia (–0.23 [– 0.58,0.12]), A/Hong Kong (–0.06 [–0.26,0.14]), and A/Singapore (–0.19 [–0.40,0.01]), although all confidence intervals overlapped the null. Two-year fold-change estimates supported these findings, with A/Cambodia (0.44 [0.29,0.65]) and A/Singapore (0.48 [0.33,0.69]) indicating significant waning, while A/Hong Kong (0.81 [0.68,1.01]) indicated minimal waning.

MN results in EPI-HK showed no evidence of monthly waning for A/H3N2/Singapore (-0.06 [-0.19, 0.07]) and for A/H3N2/Kansas (-0.05 [-0.27,0.17]). Fold-change estimates were consistent indicating minimal waning for A/H3N2/Singapore (0.79 [0.71,0.88]) and A/H3N2/Kansas (0.80 [0.57,1.09]).

#### Influenza B

In HIVE cohort, HAI monthly waning occurred for B/Colorado (–0.08 [–0.22,0.06]), while B/Phuket titers remained stable (0.03 [–0.14,0.20]). Fold-change estimates indicated no evidence of waning for B/Phuket (1.29 [1.07,1.62]) and stable antibody levels for B/Colorado (0.99 [0.78,1.22]), indicating no waning for both strains.

In EPI-HK, HAI monthly waning estimates suggested stable antibody levels for B/Phuket (0.07 [– 0.09,0.22]) and a non-significant decline for B/Washington (–0.16 [–0.65,0.349]). Fold-change estimates were consistent with these patterns, with no evidence of waning for B/Phuket (1.30 [1.15,1.44]) and B/Washington (0.53 [0.29,0.90]) indicating moderate waning.

## Discussion

In this study, we evaluated longitudinal antibody changes in two geographically distinct cohorts during a period of reduced influenza transmission. Overall, baseline antibody levels were consistently higher in HIVE than in EPI-HK across both HAI and MN assays. Antibody waning was heterogeneous across strains, with H1N1 showing moderate to significant waning in HIVE and more variable patterns in EPI-HK, while H3N2 antibody levels remained largely stable in HIVE but showed strain-specific waning in EPI-HK, particularly for A/Cambodia and A/Singapore. Influenza B antibody levels were generally stable in both cohorts, with the exception of moderate waning for B/Washington in EPI-HK. Results from MN assays were broadly consistent with HAI findings and provided additional evidence of waning for selected strains.

Cross-laboratory comparisons showed generally consistent agreement in HAI measurements between University of Michigan and University of Hong Kong laboratories, supporting the overall reproducibility of serologic testing across sites. Agreement assessed using kappa statistics varied by strain, with higher concordance observed for H1N1 and more limited agreement for certain H3N2 strains. Similarly, agreement between HAI and MN assays was moderate for H1N1 but lower for H3N2. The reduced agreement between HAI and MN for H3N2 is consistent with prior observations in literature and highlights known differences in assay performance across influenza subtypes^14,18^.

HIVE and EPI-HK represent two regions with both historical and contemporary differences in both influenza virus circulation, public health measures and vaccine distribution. These factors are clearly reflected in our observation of distinct levels of population immunity prior to the pandemic, as measured by baseline, pre-pandemic influenza antibody levels. Different vaccine policies and levels of uptake, as well as distinct pre-pandemic circulation patterns, likely shaped the baseline immunity in these populations and influenced their responses during the pandemic^19–22^. Among two study locations, HIVE participants generally had higher pre-pandemic vaccine coverage and documented prior infections, while EPI-HK participants included a larger proportion of individuals with no recent influenza vaccination history. These differences continued after the re-emergence of influenza. H3N2 strains (clade: 3C.2a1b.2a.2.) circulated extensively in Michigan during 2022–2023 season^23^, but were largely absent in Hong Kong, where masking and other mitigation strategies remained in place for a longer duration^7,24,25^.

These findings provide important insight into the immunological landscape of influenza during and following the COVID-19 pandemic. A major concern raised early in pandemic was that the near-complete absence of influenza virus circulation, driven by mitigation interventions, would result in widespread population-level immunity gaps and lead to more severe epidemics upon the resurgence of virus^2,26,27^. While influenza transmission did re-emerge in many regions after mitigation measures were relaxed^5,24,26^, our results suggest that individual-level antibody waning was heterogeneous. We observed moderate to significant waning for selected H1N1 strains, while H3N2 and influenza B responses were more variable, with evidence of waning for some strains but stability for others. Notably, for H3N2, the predominant circulating subtype in post-pandemic period, antibody levels remained largely stable in HIVE but showed strain-specific waning in EPI-HK, particularly for A/Cambodia and A/Singapore. These mixed patterns indicate that the resurgence of influenza was unlikely to be driven by rapid declines in homologous antibody levels alone. Instead, population susceptibility likely reflects a combination of pre-existing immune history, individual differences in maintenance of memory cells across strains, and possibly small regional differences in viruses^28,29^. Relatively limited antigenic drift occurred during the pandemic, and the consistency in these strains may have contributed to maintaining population immunity upon re-emergence. While prior studies have emphasized antibody waning among vaccinated individuals after peak levels have occurred at 3-4 weeks after vaccination and waning over 6-12 months after that peak^30–32^, our findings suggest that in the absence of substantial antigenic change, antibody responses may potentially remain relatively stable over longer time scales, although this stability varies by strain and population.

While we detected no substantial waning for some strains using functional anti-HA antibodies levels based on HAI assays, we cannot exclude possibility that other immune mechanisms, such as reductions in anti-neuraminidase antibodies, mucosal immunity, or T cell memory, which could contribute to the observed epidemiological patterns in both regions. Additionally, a proportion of participants experienced rising antibody titers over study period, suggesting undocumented natural exposures. This is likely due to more relaxed and shorter-duration mitigation measures in Michigan compared to Hong Kong, especially during Omicron period.

Understanding dynamics of influenza antibodies at population level is critical for informing vaccination strategies and projecting disease burden, particularly during periods of minimal antigenic drift or shift when long-term immune durability is difficult to estimate. Although short-term antibody waning following vaccination or infection has been well characterized in pre-pandemic studies^30–33^, evidence on longer-term antibody trajectories in the absence of repeated exposure remains limited. This study addresses this gap by leveraging prospective longitudinal cohorts with detailed individual-level data on vaccination history, infection surveillance, and timing of serum collection, allowing for robust estimation of antibody dynamics over multiple years. In addition, inclusion of two geographically and epidemiologically distinct populations provides a unique opportunity to evaluate how differences in vaccination coverage, prior exposure, circulating strains, and demographic factors shape patterns of antibody waning across settings. Several limitations should be considered. We were unable to comprehensively capture infection history prior to baseline sample in EPI-HK cohort, which may limit our ability to fully account for prior natural exposures, and HAI-based measures may not fully capture the breadth of immune responses relevant to protection, including non-neutralizing or cellular immunity.

In conclusion, we observed generally modest and strain-specific waning of influenza antibody titers at individual level during pandemic period in both U.S. and Hong Kong. Across both HAI and MN assays, most antibody levels remained relatively stable, although statistically significant declines were detected for selected H1N1 and H3N2 strains, particularly in EPI-HK cohort. The magnitude of waning was small and heterogeneous across strains and assays, suggesting gradual immune decay rather than rapid loss of protection. Taken together, these findings do not provide strong evidence for substantial population-level waning of influenza immunity across all subtypes and strains during the period of suppressed viral circulation. However, the presence of strain-specific declines indicates that some degree of waning did occur, and the implications for susceptibility may vary by subtype and setting. Further research is needed to clarify how baseline antibody levels, prior infection history, and vaccination interact to shape post-pandemic immunity and to determine whether gradual antibody declines translate into meaningful changes in clinical protection. Regional differences in strain circulation, public health measures, and vaccine uptake continue to offer valuable insight into how these combined factors influence vulnerability to re-emerging influenza strains.

### Ethical approval

The Household Influenza Vaccine Evaluation (HIVE) study: All adult household members provided informed consent for themselves and their children; children aged 7 years and older provided assent to participation. The study was approved by the University of Michigan Medical School Institutional Review Board (HUM00118900; HUM00198212). The Evaluating Population Immunity in Hong Kong (EPI-HK) study: Participants aged 18 years or above provided written informed. For participants aged 11-17 years, proxy written consent were obtained from parents or legal guardians in addition to their own written assent. For participants aged 0-10 years, proxy written consent from parents or legal guardians was obtained. The study was approved by the University of Hong Kong Hospital Authority Hong Kong West Cluster Institutional Review Board (UW19-720).

## Data Availability

The data used in this study can be made available upon request. Due to Institutional Review Board (IRB) regulations, data access is controlled. Per the guidelines of the Centers for Excellence in Influenza Research and Response (CEIRR) Network, individuals seeking access must complete a data and specimen collaboration form. Requests received will be reviewed by the study investigators.

## Acknowledgements

We would like to thank all individuals for their participation in the HIVE and the EPI-HK studies. We would like to thank Lewis Siu for preparing the virus antigens for EPI-HK study.

## Author contributions

Yanyupei Yang: conceptualization, formal analysis, methodology, software, visualization, writing – original draft, writing – reviewing and editing

Matthew Smith: data curation, data management, writing – reviewing and editing

Faith Ho: data curation, data management, visualization, writing – reviewing and editing

Rachel Truscon: laboratory testing, methodology, writing – reviewing and editing

Nancy H. L. Leung: conceptualization, writing – reviewing and editing

Lisa Touyon: laboratory testing, writing – reviewing and editing

Amanda Schaefbauer: laboratory testing, methodology, writing – reviewing and editing

Amy Callear: data curation, data management, writing – reviewing and editing

Elie-Tino Godonou: data curation, data management, writing – reviewing and editing

Christopher N. Blair: laboratory testing, methodology, writing – reviewing and editing

Arnold S. Monto: conceptualization, writing – reviewing and editing

Adams S. Lauring: conceptualization, methodology, writing – reviewing and editing

Benjamin J. Cowling: funding acquisition, conceptualization, methodology, writing – reviewing and editing

Sook-San Wong: funding acquisition, conceptualization, methodology, writing – reviewing and editing

Emily T. Martin: funding acquisition, conceptualization, methodology, writing – reviewing and editing

## Disclaimer

The content is solely the responsibility of the authors and does not necessarily represent the official views of the funding agencies.

