## Supplemental materials for "A prospective, comparative cohort analysis of influenza antibody waning in Michigan and Hong Kong during periods of low influenza circulation"

### Supplementary materials

#### Supplementary methods

##### Eligibility criteria

For this analysis, the Household Influenza Vaccine Evaluation (HIVE) study eligible participants met the following criteria:

1. Participated in the study from fall 2020 to fall 2022.
2. No self-reported or influenza vaccination records from fall 2020 to fall 2022.
3. Availability of longitudinal serological samples (≥1 blood sample/year) across four collection periods: pre-season 2020-21, post-season 2020-21, pre-season 2021-22, and post-season 2021-22.

For this analysis, the Evaluating Population Immunity in Hong Kong (EPI-HK) study eligible participants met the following criteria:

1. Participated in the study from fall 2020 to fall 2022.
2. No self-reported or influenza vaccination records from fall 2020 to fall 2022.
3. Provide longitudinal blood samples collected approximately every six months for four periods from fall 2020 (July–December 2020) to fall 2022 (July–December 2022).
4. Be randomly selected from six age groups (0–17, 18–29, 30–44, 45–54, 55–64, 65–74 years), with 10 participants per age group.

##### HAI testing

The hemagglutination inhibition (HAI) assay was performed to measure serum antibodies that block hemagglutinin (HA) receptor binding. Prior to HAI testing, all serum specimens were treated overnight with receptor-destroying enzyme and subsequently heat-inactivated to eliminate nonspecific inhibition. In addition, all sera were adsorbed with turkey red blood cells to remove nonspecific agglutinins.

Ten serial two-fold dilutions were prepared for each serum specimen, starting with an initial dilution of 1:10 and ending at 1:5120. Diluted sera were distributed into 96-well V-bottom microtiter plates. The serum dilutions were then incubated with standardized concentrations of either egg- or cell-grown antigens (4 HA units per 25 μL). After serum and antigen incubation, standardized turkey red blood cells were added to each well and allowed to settle. The strain-specific HAI antibody titers were determined for each time point and each individual as the reciprocal of the highest serum dilution that completely inhibited hemagglutination (e.g., a 1:40 dilution was recorded as a titer of 40). Titers below the assay's limit of detection (<1:10) were assigned a value of 5, representing half the lower detection threshold. Titers exceeding the highest dilution tested (>1:5120) were assigned a value of 10240, corresponding to twice the upper detection limit.

Antigen target selection and preparations differed slightly between cohorts. For the HIVE cohort, the antigens reflected the vaccine composition in the 2019–2020 Northern Hemisphere influenza vaccine (Table S2). In order to obtain the required volume and hemagglutinin (HA) titer of the cell targets for the HIVE HAI, seed viruses were sourced from the Influenza Reagent Resource (IRR) and propagated at the University of Michigan in MDCK-SIAT1 cells. One cell target, B/Colorado, was sourced from IRR inactivated. All egg targets were sourced from IRR as inactivated antigens. Due to biosafety concerns and the high risk associated with handling a live virus that was no longer circulating, B/Phuket (Yamagata lineage) was not tested using live cell-cultured virus. An inactivated cell-grown virus was unavailable. For A/Singapore, a cell seed virus was not available for propagation, the egg-grown seed virus did not yield an HA titer with turkey red blood cells after passage into MDCK-SIAT1 cells.

For the EPI-HK cohort, the antigens reflected the vaccine composition of the 2015-2023 Northern Hemisphere influenza vaccine (Table S2). Viruses were originally sourced from WHO Collaborating Center in Melbourne or St. Jude’s Children Research Hospital and subsequently minimally propagated in-house accordingly. All virus stocks were sequenced and confirmed to have no significant antigenic changes prior to use. Handling of antigens (egg-grown or cell-grown) and virus inactivation protocols were performed according to standard procedures^1^.

##### Microneutralization testing (MN)

Assay targets were generated as recombinant 6+2 influenza A viruses using standard reverse
genetics. Briefly, viral stocks of A/Michigan/45/2015 (H1N1), A/Idaho/07/2018 (H1N1),
A/Singapore/INFIMH-16-0019/2016 (H3N2), A/Kansas/14/2017 (H3N2) were obtained from the
International Reagent Resource (IRR), and segment 4 (HA) and segment 6 (NA) were amplified
by RT-PCR as in^2^ and cloned into pHW2000^3^. Recombinant
luciferase-expressing influenza viruses with the corresponding HA and NA proteins were
rescued following transfection of MDCK-SIAT1 and HEK293T co-cultures with TransIT-LT1 (MIRUS
#MIR2300) and the following plasmids: PASTN-PA-NanoLuc (a gift from
Andrew Mehle, University of Wisconsin^4^), strain specific pHW184-HA, strain specific pHW186-
NA, pHW181-PB2, pHW182-PB1, pHW185-NP, pHW187-M, pHW188-NS. The PB2, PB2, NP, M,
and NS segments were on A/WSN/1933 plasmid backbones. Transfection supernatants were
passaged once on MDCK-SIAT1 cells, these passage 1 stocks were used in the
microneutralization assay.

Luciferase microneutralization (LMN) assays were performed as previously described^5^. Briefly, human (positive) control serum from a vaccinated donor, goat (negative)
control serum from IRR (FR-1377), and subject sera were heat inactivated at 56˚ C for 30 min
and two-fold, serially diluted across columns of a 96 well plate. For each target virus, the P1
stock was diluted to a concentration of 2x103 TCID50/mL and 50μL (100 infectious units) were
added to all the serum dilution wells. A back titer series was created by adding 50μL (100
infectious units) to the first well in a column and then performing two-fold serial dilutions down
to the last well. After a one hour incubation at 37˚C, 100μL MDCK-SIAT1 cells at a
concentration of 1.5x105 cells/mL were added to each well. The plate was then incubated for
18h at 37˚ C. After incubation, the supernatants were aspirated from the wells and 50 μL of a
1:8000 concentration of ViviRen (Promega, #E6491) in viral media were added to each well. The
plate was incubated at room temperature for 3 min, and luminescence was measured in a
Synergy HTX multi-mode luminometer using a gain of 160. All the steps involving ViviRen were
performed in a darkened room.

##### Statistical methods

To evaluate the waning of antibody titers over time, we modeled the association between hemagglutination inhibition (HAI) titers and time since baseline using three approaches: (1) linear mixed-effects models, (2) exponential decay models, and (3) natural cubic spline models. All analyses were conducted on log₂-transformed titer values.

1. Linear Mixed-Effects Model (LMM)

The linear mixed-effects model assumes a constant change in antibody titers over time. For participant i at time point j, the model is specified as:

log₂(titer_ij_) = β₀ + β₁ × time_ij_ + β₂ × age_ij_ + β₃ × baseline_titer_i_ + b₀_i_ + ε_ij_

- titer_ij_: observed titer for participant i at time j

##### - time_ij_: time since baseline (in months)

- age_ij_: participant age at baseline (continuous)

- baseline_titer_i: participant’s first titer measurement

- b₀_i_: random intercept for participant i

- ε_ij_: residual error

2. Exponential Decay Model

This model assumes a power-law decline in titers over time, where antibody waning follows a non-linear but monotonic pattern. The model is specified as:

log₂(titer_ij_) = β₀ + β₁ ×log₂(time_ij_)+ β₂ × age_ij_ + β₃ × baseline_titer_i_ + b₀_i_ + ε_ij_

3. Natural Cubic Spline Model

To allow for potential non-linear waning patterns, we used a natural cubic spline with three knots placed at quantiles of the time variable. The model is specified as:

log₂(titer_ij_) = β₀ + f(time_ij_) + β₂ × age_ij_ + β₃ × baseline_titer_i_ + b₀_i_ + ε_ij_

Where f(·) is a natural cubic spline basis function of time. Knots were placed at the 10th, 50th, and 90th percentiles of observed follow-up times.

Model Fitting and Selection

Each model was fitted separately for each antigen strain. Both univariate and multivariate versions were assessed. Univariate models included only time as a fixed effect; multivariate models adjusted for age and baseline titer. Model fit was compared using Akaike Information Criterion (AIC) and Bayesian Information Criterion (BIC); models with the lowest AIC and BIC values were selected as best fitting.

All statistical analyses were performed using R version 4.3.1. Mixed-effects models were implemented using the “lme4” package, and spline models using the “splines” package.

Table S1: Participants and serological sample

| **region** | **Age group** | **Number of Participants** | **Number of Samples** |
| --- | --- | --- | --- |
| Michigan | 0-17 | 21 | 62 |
| Michigan | 18-29 | 6 | 16 |
| Michigan | 30-44 | 18 | 54 |
| Michigan | 45-54 | 11 | 38 |
| Michigan | 55-64 | 1 | 3 |
| HK | 0-17 | 10 | 40 |
| HK | 18-29 | 10 | 44 |
| HK | 30-44 | 10 | 42 |
| HK | 45-54 | 10 | 49 |
| HK | 55-64 | 11 | 47 |
| HK | 65+ | 9 | 37 |
| HK additional | 0-17 | 2 | 8 |
| HK additional | 18-29 | 30 | 120 |
| HK additional | 30-44 | 30 | 120 |
| HK additional | 45-54 | 30 | 120 |
| HK additional | 55-64 | 31 | 123 |
| HK additional | 65+ | 29 | 115 |

Table S2: Influenza virus antigens used for hemagglutination inhibition (HAI) assays in the HIVE and EPI-HK cohorts, 2020–2023

| **Target antigens** | **HIVE antigen** | **EPI-HK antigen** |
| --- | --- | --- |
| **Egg antigens** | H1N1pdm09 A/Brisbane/02/2018  H3N2 A/Singapore/INFIMH-16-0019/2016  Yamagata B/Phuket/3072/2013 | H1N1pdm09  A/Michigan/45/2015  H1N1pdm09 A/Brisbane/02/2018  H1N1 A/Victoria/2570/2019  H3N2 A/Hong Kong/4801/2014  H3N2 A/Singapore/INFIMH-0019-2016  H3N2 A/Kansas/14/2017 (2019-2020)  H3N2 A/Cambodia/e0826360/2020 (2021-2022)  Yamagata B/Phuket/3072/2013  Victoria B/Washington/02/2019 |
| **Cell antigens** | H1N1pdm09  A/Michigan/45/2015  A/Idaho/07/2018  H3N2 A/Kansas/14/2017  Victoria B/Colorado/06/2017 | H1N1 A/Hawaii/70/2019 |

Figure S1: Geometric mean titer at baseline and over time by region

A. Baseline hemagglutination inhibition (HAI) antibody titers by cohort. Violin plots show the distribution of baseline HAI titers for each strain, with overlaid boxplots indicating the median and interquartile range and individual points representing participants. Titers are displayed on a log₂ scale and labeled as reciprocal antibody titers. P-values were calculated using the Wilcoxon rank-sum test comparing baseline titers between cohorts for each strain independently. HIVE, Household Influenza Vaccine Evaluation cohort; EPI-HK, Evaluating Population Immunity in Hong Kong cohort.

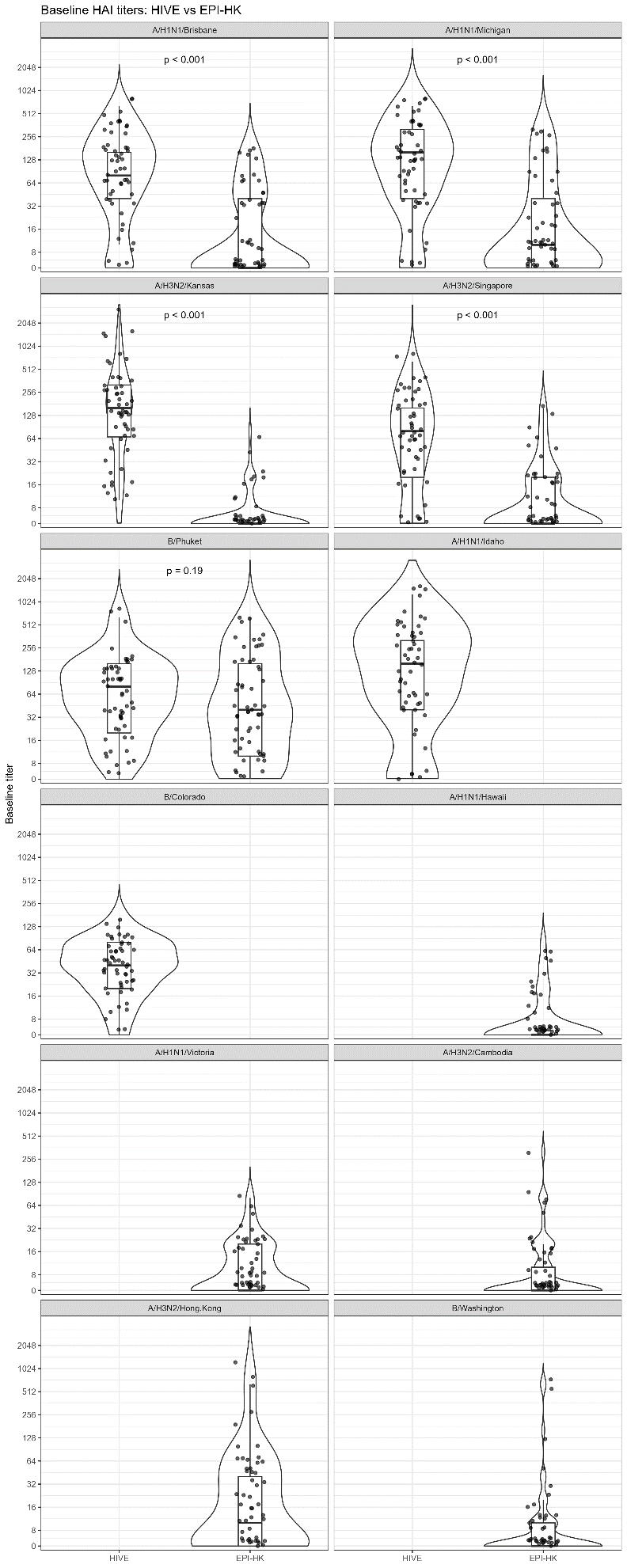

B. Baseline microneutralization (MN) antibody titers between the HIVE and EPI-HK cohorts. Violin plots show the distribution of baseline MN titers for each strain, with overlaid boxplots indicating the median and interquartile range and individual points representing participants. Titers are displayed on a log₂ scale and labeled as reciprocal antibody titers. P-values were calculated using the Wilcoxon rank-sum test comparing baseline titers between cohorts for each strain independently. HIVE, Household Influenza Vaccine Evaluation cohort; EPI-HK, Evaluating Population Immunity in Hong Kong cohort.

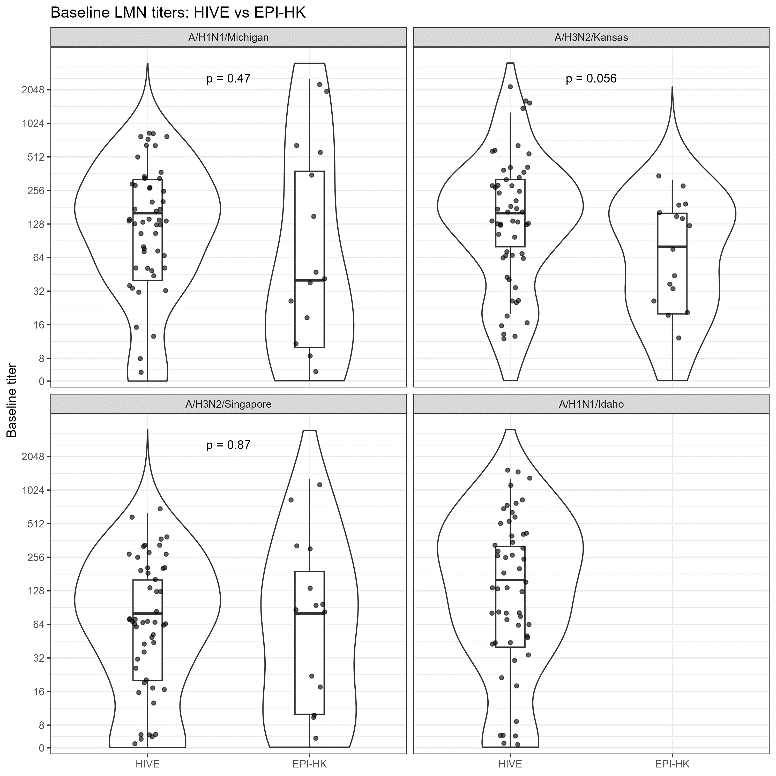

C. HAI Geometric mean antibody titers (GMTs) over time by cohorts. Points represent strain-specific geometric mean titers (GMTs), and error bars indicate 95% confidence intervals. GMTs were estimated from serum specimens collected longitudinally from participants without documented influenza infection or vaccination during the study period. Panels are stratified by influenza strain, and values are shown separately for the HIVE and EPI-HK cohorts.

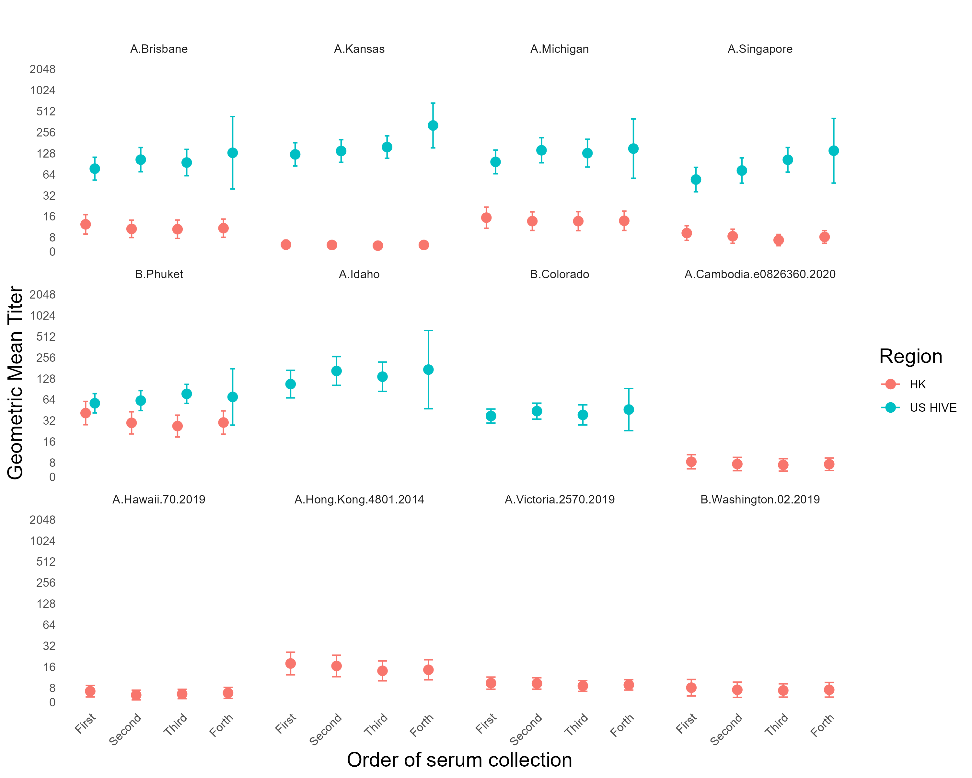

D. HAI Geometric mean antibody titers (GMTs) by serum collection period, age group, and cohort. Points represent strain-specific geometric mean titers (GMTs), and error bars indicate 95% confidence intervals. GMTs were calculated separately by serum collection period, age category, and cohort among participants without documented influenza infection or vaccination during the study period. Colors indicate cohort (HIVE, Michigan; EPI-HK, Hong Kong), and point shapes indicate age category. Panels are stratified by influenza strain.

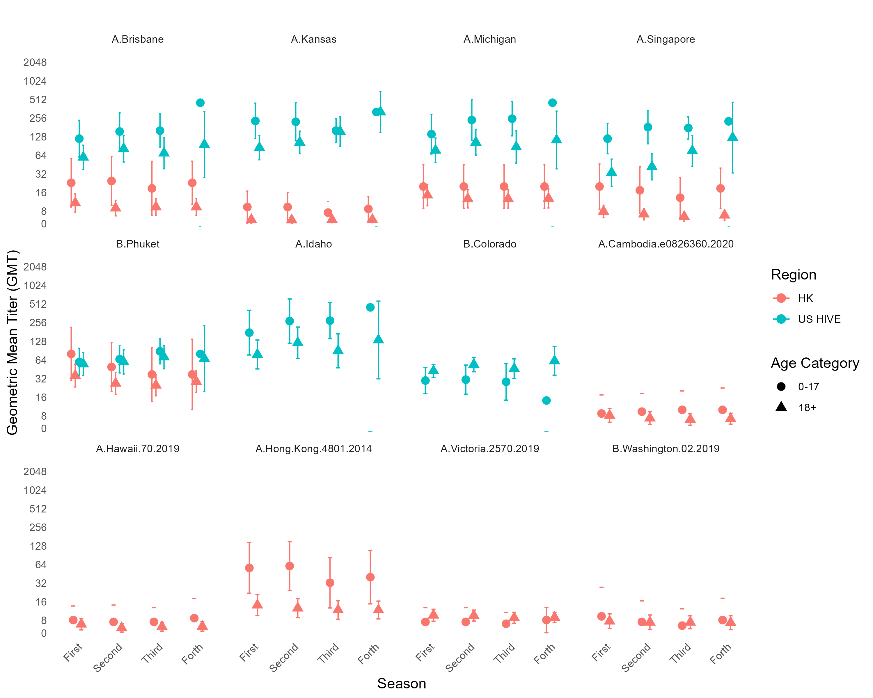

Figure S2: Inter-laboratory comparison of hemagglutination inhibition (HAI) antibody titers measured at the University of Hong Kong and University of Michigan laboratories. Scatterplots compare strain-specific HAI titers measured in the University of Hong Kong laboratory and the University of Michigan laboratory among a subset of EPI-HK serum specimens tested in both laboratories. Each point represents an individual serum specimen. The color of the point represents the participants. The dashed line indicates perfect agreement between laboratories (y = x). Antibody titers are displayed on a common log₂-scaled axis and labeled as reciprocal titers. Pearson correlation coefficients (r) quantify agreement in continuous titer measurements, while Cohen’s kappa statistics (κ) assess agreement in seroprotection status defined as HAI titer ≥1:40 versus <1:40. Values of r, κ, and the number of paired specimens (N) are shown within each panel.

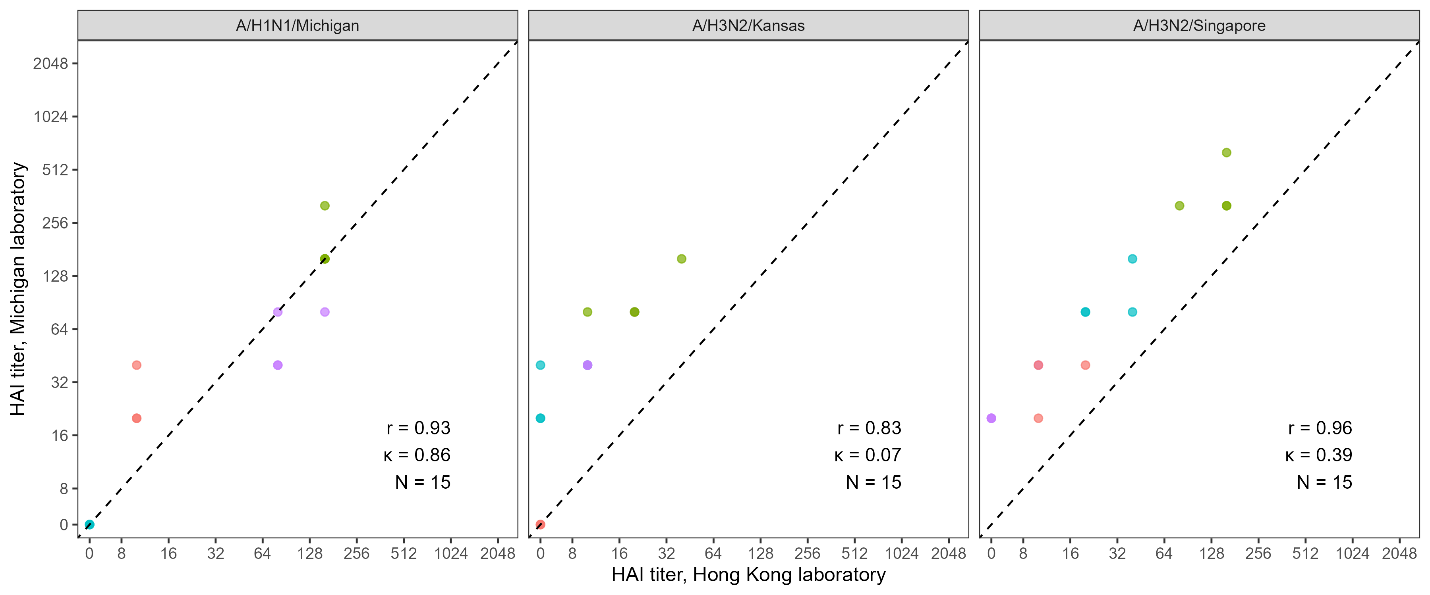

Figure S3: Comparison of hemagglutination inhibition (HAI) and microneutralization (MN) antibody titers among HIVE serum specimens. Scatterplots compare strain-specific HAI and MN antibody titers measured from the same serum specimens in the HIVE cohort. Each point represents an individual serum specimen. The color of the point represents the participants. The dashed line indicates perfect agreement between assay platforms (y = x). Antibody titers are displayed on a common log₂-scaled axis and labeled as reciprocal titers. Pearson correlation coefficients (r) quantify agreement in continuous antibody measurements, while Cohen’s kappa statistics (κ) assess agreement in seroprotection status defined as antibody titers ≥1:40 versus <1:40. Values of r, κ, and the number of paired specimens (N) are shown within each panel.

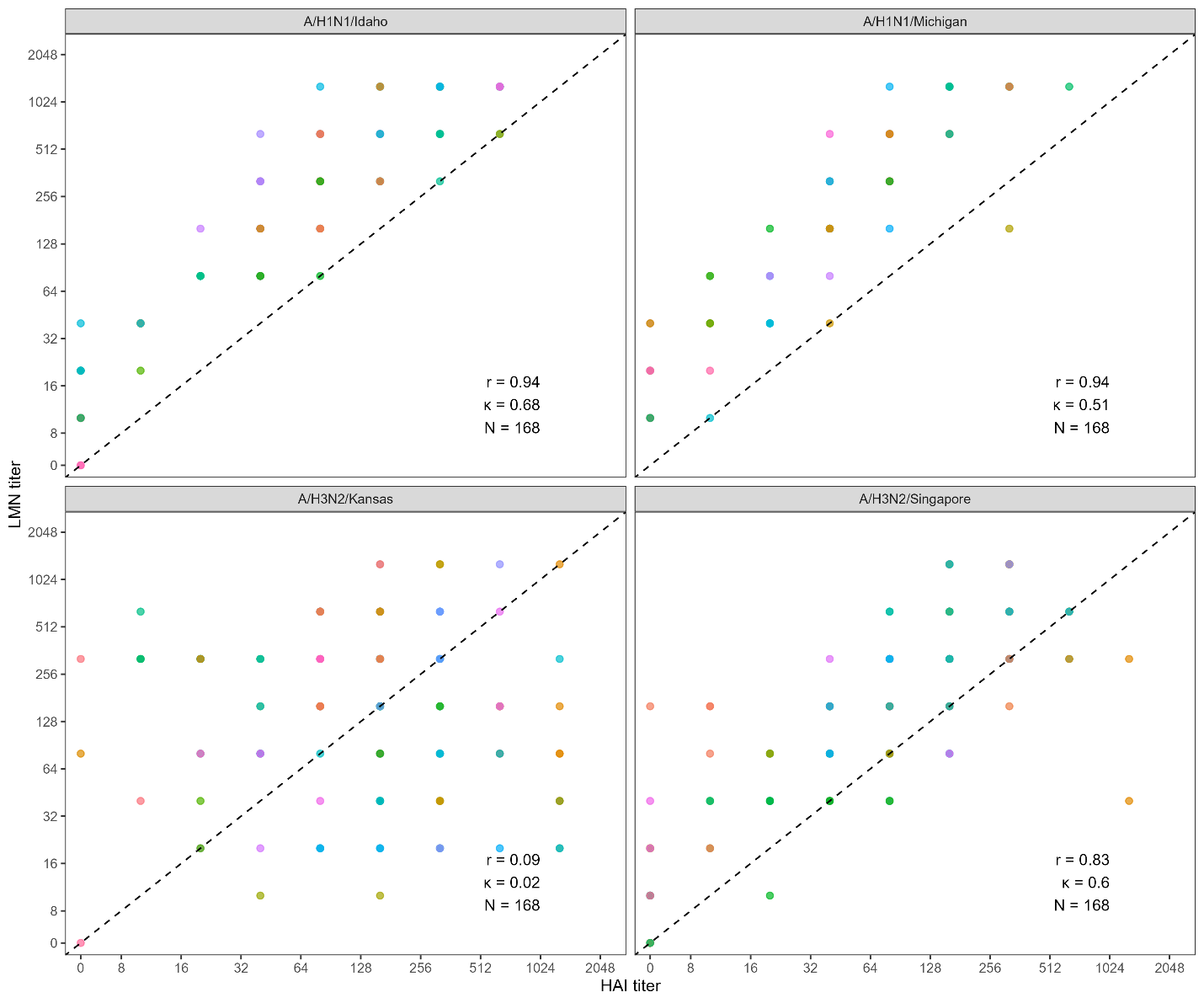

Figure S4: Comparison of hemagglutination inhibition (HAI) and microneutralization (MN) antibody titers among EPI-HK serum specimens tested at the University of Michigan laboratory. Scatterplots compare strain-specific HAI and MN antibody titers measured from the same serum specimens. Each point represents an individual serum specimen. The color of the point represents the participants. The dashed line indicates perfect agreement between assay platforms (y = x). Antibody titers are displayed on a common log₂-scaled axis and labeled as reciprocal titers. Pearson correlation coefficients (r) quantify agreement in continuous antibody measurements, while Cohen’s kappa statistics (κ) assess agreement in seroprotection status defined as antibody titers ≥1:40 versus <1:40. Values of r, κ, and the number of paired specimens (N) are shown within each panel.

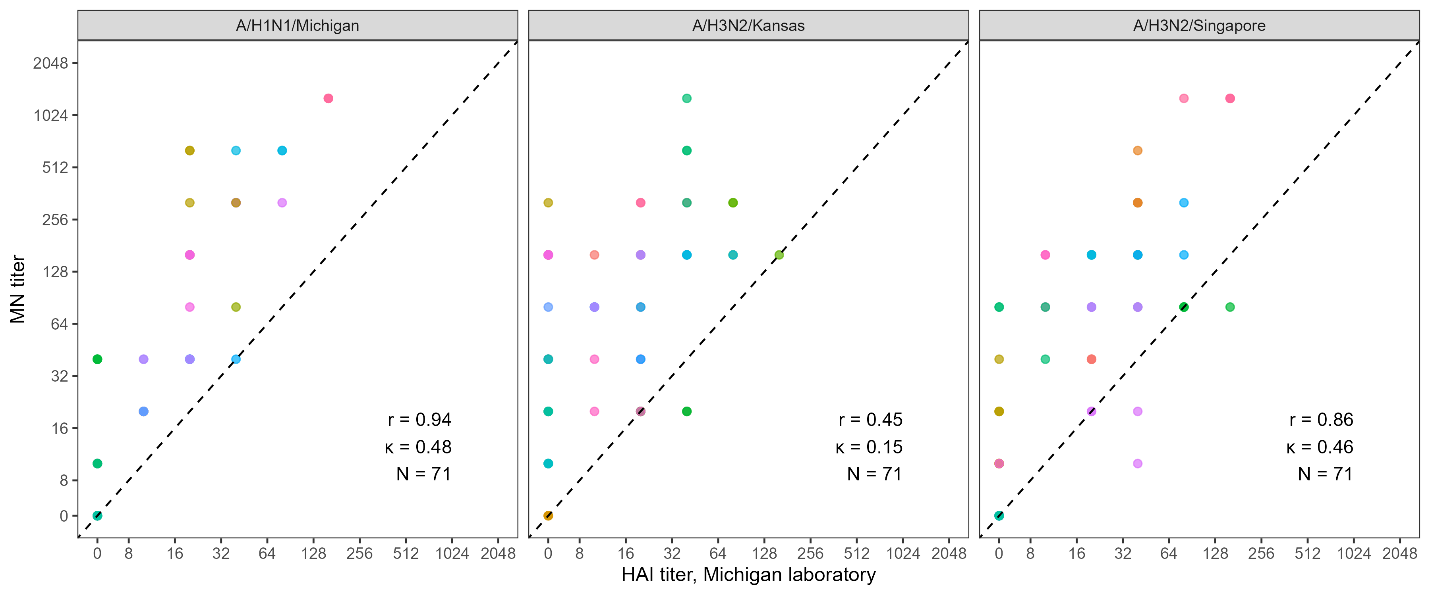

Figure S5: Predicted longitudinal trajectories of HAI antibody titers among participants with baseline seroprotective antibody levels (HAI titer ≥1:40), stratified by age group. Model-predicted HAI antibody titers were estimated from the best-fitting multivariable waning models, adjusted for age and baseline antibody titer. Curves represent predicted antibody trajectories over time for participants with baseline HAI titers ≥1:40, with separate lines shown for each age group.

A. HIVE cohort (Michigan). Predicted HAI antibody trajectories among participants with baseline HAI titers ≥1:40.

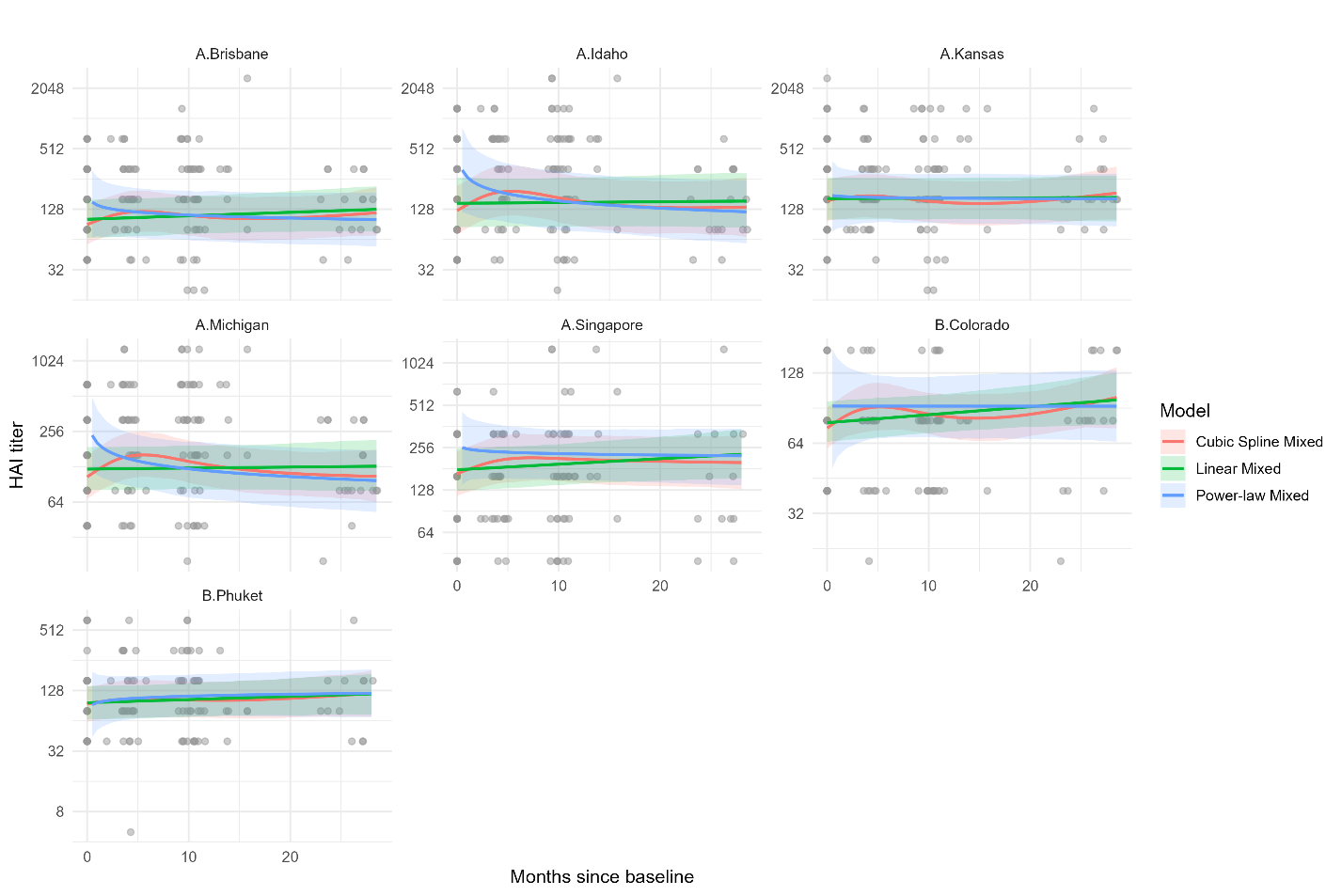

B. EPI-HK cohort (Hong Kong). Predicted HAI antibody trajectories among participants with baseline HAI titers ≥1:40.

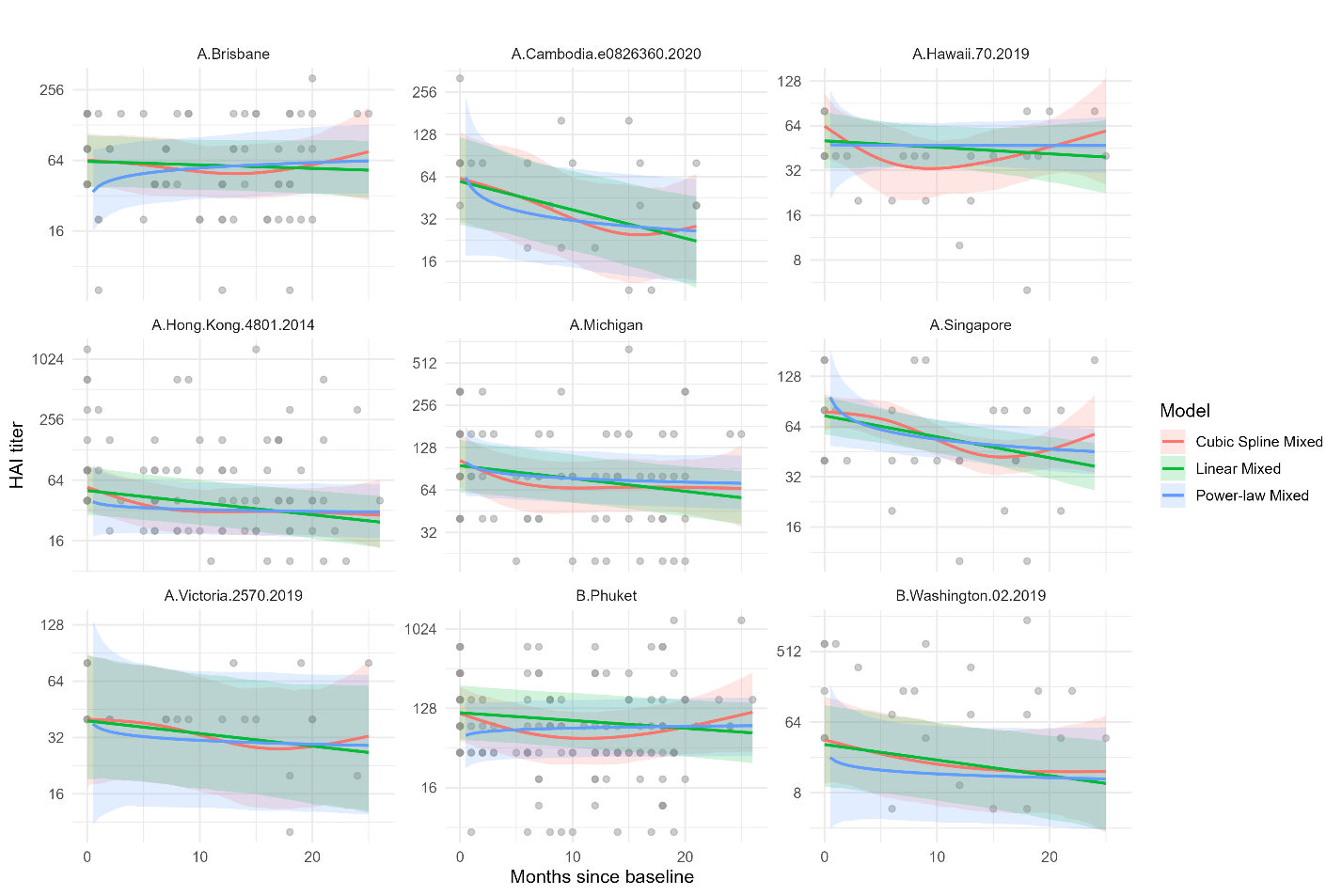

Table S3: HAI GMT and fold change by age and cohorts

| **cohort** | **Test** | **strain** | **antigen** | All participants | | | | <18 years participants | | | 18 + years participants | | |
| --- | --- | --- | --- | --- | --- | --- | --- | --- | --- | --- | --- | --- | --- |
|  |  |  |  | **First sample GMT (95%CI)** | **Fourth sample GMT (95% CI)** | **Overall fold change (95% CI)** | **Increased titer participants (%)** | **First sample GMT (95%CI)** | **Fourth sample GMT (95% CI)** | **Overall fold change (95% CI)** | **First sample GMT (95%CI)** | **Forth sample GMT (95% CI)** | **Overall fold change (95% CI)** |
| HIVE | HAI | H1N1 | A.Brisbane | 77.08 (52.87, 112.38) | 129.96 (39.42, 428.48) | 1.69 (0.48, 5.98) | 12 (21.4%) | 118.88 (60.17, 234.86) | 452.55 (0, 1652102041060.23) | 3.81 (0, 20469240147.81 ) | 59.44 (37.96, 93.07) | 95.14 (27.83, 325.25) | 1.6 (0.42, 6.08 ) |
| HIVE | HAI | H1N1 | A.Idaho | 106.35 (67.51, 167.52) | 171.48 (47.21, 622.86) | 1.61 (0.4, 6.46) | 15 (26.8%) | 176.65 (76.99, 405.33) | 452.55 (0, 1652102041060.23) | 2.56 (0, 16598214655.82 ) | 78.43 (45.84, 134.19) | 134.54 (31.73, 570.47) | 1.72 (0.36, 8.28 ) |
| HIVE | HAI | H1N1 | A.Michigan | 96.32 (65.1, 142.52) | 149.29 (56.23, 396.35) | 1.55 (0.53, 4.53) | 16 (28.6%) | 140.21 (67.07, 293.11) | 452.55 (0, 247222566.75) | 3.23 (0, 3705544.32 ) | 76.89 (48.63, 121.58) | 113.14 (38.69, 330.83) | 1.47 (0.44, 4.88 ) |
| HIVE | HAI | H3N2 | A.Kansas | 123.38 (84.05, 181.1) | 320 (152.8, 670.14) | 2.59 (1.1, 6.11) | 8 (14.3%) | 230.04 (119.02, 444.62) | 320 (0, 14290502215.15) | 1.39 (0, 97577865.49 ) | 84.9 (54.2, 132.97) | 320 (149.84, 683.38) | 3.77 (1.5, 9.47 ) |
| HIVE | HAI | H3N2 | A.Singapore | 53.84 (35.97, 80.59) | 139.29 (47.97, 404.47) | 2.59 (0.81, 8.25) | 13 (23.2%) | 118.88 (67.86, 208.25) | 226.27 (0, 123611283.37) | 1.9 (0, 1605213.74 ) | 33.47 (20.21, 55.44) | 123.38 (33.23, 458.11) | 3.69 (0.88, 15.52 ) |
| HIVE | HAI | B.Victoria | B.Colorado | 37.14 (29.47, 46.8) | 45.95 (22.97, 91.92) | 1.24 (0.59, 2.6) | 12 (21.4%) | 29.72 (18.31, 48.24) | 14.14 (0, 7725705.21) | 0.48 (0, 359964.51 ) | 42.45 (33.37, 54) | 61.69 (36.28, 104.9) | 1.45 (0.8, 2.65 ) |
| HIVE | HAI | B.Yamagata | B.Phuket | 56.57 (41.14, 77.79) | 69.64 (27.5, 176.36) | 1.23 (0.45, 3.33) | 9 (16.1%) | 59.44 (35.89, 98.45) | 80 (80, 80) | 1.35 (0.06, 29.09 ) | 54.91 (35.74, 84.38) | 67.27 (19.68, 229.99) | 1.23 (0.32, 4.62 ) |
| EPI-HK | HAI | H1N1 | A.Brisbane | 12.31 (8.95, 16.93) | 10.84 (8.03, 14.64) | 0.88 (0.57, 1.36) | 4(6.7%) | 22.97 (9.35, 56.46) | 22.97 (10.29, 51.28) | 1 (0.3, 3.34) | 10.87 (7.72, 15.29) | 9.33 (6.78, 12.84) | 0.86 (0.54, 1.37) |
| EPI-HK | HAI | H1N1 | A.Hawaii | 7.15 (5.91, 8.66) | 6.75 (5.63, 8.1) | 0.94 (0.73, 1.23) | 4(6.7%) | 8.12 (4.8, 13.73) | 8.71 (4.19, 18.1) | 1.07 (0.44, 2.64) | 6.97 (5.64, 8.62) | 6.42 (5.38, 7.66) | 0.92 (0.7, 1.21) |
| EPI-HK | HAI | H1N1 | A.Michigan | 15.33 (10.82, 21.74) | 13.82 (10.05, 19.01) | 0.90 (0.56, 1.45) | 7(11.7%) | 20 (8.9, 44.95) | 20 (8.9, 44.95) | 1 (0.32, 3.14) | 14.54 (9.78, 21.61) | 12.83 (9, 18.31) | 0.88 (0.52, 1.5) |
| EPI-HK | HAI | H1N1 | A.Victoria | 9.33 (7.64, 11.39) | 8.81 (7.43, 10.44) | 0.94 (0.73, 1.23) | 10(16.7%) | 7.58 (4.45, 12.91) | 8.12 (5.07, 13) | 1.07 (0.53, 2.18) | 9.73 (7.79, 12.14) | 8.95 (7.41, 10.8) | 0.92 (0.69, 1.23) |
| EPI-HK | HAI | H3N2 | A.Cambodia | 8.22 (6.51, 10.38) | 7.58 (6.17, 9.32) | 0.92 (0.68, 1.26) | 3(5%) | 8.71 (4.35, 17.42) | 10 (4.45, 22.47) | 1.15 (0.4, 3.34) | 8.12 (6.29, 10.49) | 7.17 (5.85, 8.79) | 0.88 (0.64, 1.22) |
| EPI-HK | HAI | H3N2 | A.Hong.Kong | 17.82 (12.27, 25.88) | 14.47 (10.4, 20.14) | 0.81 (0.49, 1.34) | 4(6.7%) | 56.57 (22.05, 145.14) | 40 (14.84, 107.83) | 0.71 (0.18, 2.78) | 14.14 (9.59, 20.85) | 11.81 (8.47, 16.46) | 0.84 (0.5, 1.39) |
| EPI-HK | HAI | H3N2 | A.Kansas | 6.3 (5.42, 7.32) | 6.23 (5.52, 7.03) | 0.99 (0.82, 1.2) | 6(10%) | 9.33 (5.15, 16.89) | 8.71 (5.52, 13.73) | 0.93 (0.44, 1.97) | 5.82 (5.07, 6.7) | 5.82 (5.19, 6.53) | 1 (0.83, 1.2) |
| EPI-HK | HAI | H3N2 | A.Singapore | 9.22 (7.24, 11.75) | 8.12 (6.59, 10.01) | 0.88 (0.64, 1.21) | 2(3.3%) | 20 (8.61, 46.46) | 18.66 (8.77, 39.73) | 0.93 (0.3, 2.89) | 7.9 (6.27, 9.95) | 6.88 (5.73, 8.26) | 0.87 (0.65, 1.17) |
| EPI-HK | HAI | B.Yamagata | B.Phuket | 40.93 (27.83, 60.21) | 29.97 (20.42, 43.97) | 0.73 (0.42, 1.26) | 14(23.3%) | 80 (29.68, 215.66) | 37.32 (10.06, 138.44) | 0.47 (0.09, 2.41) | 35.8 (23.44, 54.69) | 28.68 (19.14, 42.97) | 0.8 (0.45, 1.44) |
| EPI-HK | HAI | B.Victoria | B.Washington | 8.03 (6.12, 10.53) | 7.49 (5.88, 9.55) | 0.93 (0.65, 1.34) | 9(15%) | 9.33 (3.16, 27.55) | 8.12 (3.61, 18.28) | 0.87 (0.23, 3.37) | 7.79 (5.95, 10.2) | 7.37 (5.69, 9.54) | 0.95 (0.65, 1.37) |
| HIVE | MN | H1N1 | A.Idaho | 399.86 (251.32, 636.19) | 452.55 (150.25, 1363.03) | 1.13 (0.33, 3.84) | 12(21%) | 735.17 (324.01, 1668.08) | 1280 (1280, 1280) | 1.74 (0.01 ,251.76) | 285.09 (162.97, 498.71) | 348.96 (89.06, 1367.28) | 1.22 (0.27 ,5.56) |
| HIVE | MN | H1N1 | A.Michigan | 656.04 (402.76, 1068.59) | 787.93 (262.91, 2361.42) | 1.2 (0.35, 4.1) | 10(18%) | 288.4 (134.74, 617.28) | 20 (20, 20) | 0.07 (0 ,7.04) | 201.59 (138.4, 293.62) | 113.14 (52.98, 241.61) | 0.56 (0.23 ,1.35) |
| HIVE | MN | H3N2 | A.Kansas | 229.09 (161.09, 325.81) | 80 (36.85, 173.69) | 0.35 (0.15, 0.83) | 10(18%) | 1194.28 (508.21, 2806.54) | 2560 (0.38, 17107585.61) | 2.14 (0 ,58900.76) | 470.32 (259.54, 852.26) | 586.88 (160.99, 2139.39) | 1.25 (0.29 ,5.41) |
| HIVE | MN | H3N2 | A.Singapore | 166.05 (108.04, 255.21) | 121.26 (51.87, 283.48) | 0.73 (0.27, 1.94) | 11(20%) | 557.15 (297.33, 1044.01) | 452.55 (5.54, 36994.69) | 0.81 (0 ,274.96) | 84.76 (54.05, 132.9) | 87.24 (35.48, 214.52) | 1.03 (0.36 ,2.92) |
| EPI-HK | MN | H1N1 | A.Michigan | 80 (23.73, 269.65) | 65.99 (22.5, 193.51) | 0.82 (0.16, 4.19) | 0(0%) | 160 (160, 160) | 126.99 (9.15, 1762.22) | 0.25 (0, 174038.67) | 55.28 (27.21, 112.29) | 52.78 (24.9, 111.86) | 0.83 (0.17, 4.1) |
| EPI-HK | MN | H3N2 | A.Kansas | 62.64 (32.97, 119) | 61.1 (31.59, 118.18) | 0.98 (0.39, 2.45) | 4(24%) | 2560 (2560, 2560) | 640 (6.72, 60907.77) | 0.79 (0, 1874.75) | 50.4 (15.69, 161.84) | 41.89 (14.08, 124.62) | 0.95 (0.34, 2.68) |
| EPI-HK | MN | H3N2 | A.Singapore | 67.96 (23.41, 197.27) | 56.57 (21.15, 151.31) | 0.83 (0.19, 3.56) | 1(6%) | 2560 (0.38, 17107585.61) | 1015.94 (13.33, 77408.49) | 0.4 (0, 2214124.98) | 41.89 (16.84, 104.22) | 31.75 (14.03, 71.83) | 0.76 (0.22, 2.58) |

Table S4: Number of participants with seroprotective antibody titers at the earliest and latest serum collection and duration of follow-up by cohort, assay, and influenza strain.

| cohort | Assay^1^ | Strain | N | Earliest titer ≥40, n (%)^2^ | Latest titer ≥40, n (%)^2^ | Days between earliest and latest serum, median (IQR)^3^ |
| --- | --- | --- | --- | --- | --- | --- |
| HIVE | HAI | A/H1N1/Brisbane | 56 | 44 (78.6%) | 43 (76.8%) | 318 (296, 474) |
| HIVE | HAI | A/H1N1/Idaho | 56 | 45 (80.4%) | 46 (82.1%) | 318 (296, 474) |
| HIVE | HAI | A/H1N1/Michigan | 56 | 47 (83.9%) | 47 (83.9%) | 318 (296, 474) |
| HIVE | HAI | A/H3N2/Kansas | 56 | 45 (80.4%) | 47 (83.9%) | 318 (296, 474) |
| HIVE | HAI | A/H3N2/Singapore | 56 | 38 (67.9%) | 41 (73.2%) | 318 (296, 474) |
| HIVE | HAI | B/Colorado | 56 | 38 (67.9%) | 39 (69.6%) | 318 (296, 474) |
| HIVE | HAI | B/Phuket | 56 | 41 (73.2%) | 44 (78.6%) | 318 (296, 474) |
| HIVE | MN | A/H1N1/Idaho | 57 | 49 (86%) | 51 (89.5%) | 319 (288, 399) |
| HIVE | MN | A/H1N1/Michigan | 57 | 52 (91.2%) | 52 (91.2%) | 319 (288, 399) |
| HIVE | MN | A/H3N2/Kansas | 57 | 53 (93%) | 46 (80.7%) | 319 (288, 399) |
| HIVE | MN | A/H3N2/Singapore | 57 | 47 (82.5%) | 49 (86%) | 319 (288, 399) |
| EPI-HK | HAI | A/H1N1/Brisbane | 60 | 18 (30%) | 12 (20%) | 549 (518, 610) |
| EPI-HK | HAI | A/H1N1/Michigan | 60 | 16 (26.7%) | 14 (23.3%) | 549 (518, 610) |
| EPI-HK | HAI | A/H3N2/Singapore | 60 | 7 (11.7%) | 5 (8.3%) | 549 (518, 610) |
| EPI-HK | HAI | A/H3N2/Kansas | 60 | 2 (3.3%) | 1 (1.7%) | 549 (518, 610) |
| EPI-HK | HAI | B/Phuket | 60 | 35 (58.3%) | 31 (51.7%) | 549 (518, 610) |
| EPI-HK | HAI | B/Washington | 60 | 5 (8.3%) | 4 (6.7%) | 549 (518, 610) |
| EPI-HK | HAI | A/H1N1/Hawaii | 60 | 5 (8.3%) | 6 (10%) | 549 (518, 610) |
| EPI-HK | HAI | A/H1N1/Victoria | 60 | 5 (8.3%) | 3 (5%) | 549 (518, 610) |
| EPI-HK | HAI | A/H3N2/Cambodia | 60 | 5 (8.3%) | 6 (10%) | 549 (518, 610) |
| EPI-HK | HAI | A/H3N2/Hong.Kong | 60 | 21 (35%) | 18 (30%) | 549 (518, 610) |
| EPI-HK | MN | A/H1N1/Michigan | 18 | 11 (61.1%) | 11 (61.1%) | 548 (525, 525) |
| EPI-HK | MN | A/H3N2/Kansas | 18 | 13 (72.2%) | 11 (61.1%) | 548 (525, 525) |
| EPI-HK | MN | A/H3N2/Singapore | 18 | 11 (61.1%) | 12 (66.7%) | 548 (525, 525) |

^1^HAI, hemagglutination inhibition assay; MN, microneutralization assay.

^2^ Values are presented as n (%) of participants with antibody titers ≥1:40 at the earliest and latest available serum collection for each strain-specific analysis.

^3^Follow-up duration is summarized as the median number of days between the earliest and latest serum specimens with interquartile range (IQR).

Table S5: Antibody waning HAI model results for HIVE

| Strain | Type | Model | Change by month (95% CI) | AIC | BIC |
| --- | --- | --- | --- | --- | --- |
| A.Brisbane | Unadjusted, all participants | Cubic Spline Mixed |  | 569.4 | 588.2 |
| A.Brisbane | Unadjusted, all participants | Linear Mixed | 0.011 [-0.005–0.027] | 577.4 | 589.9 |
| A.Brisbane | Unadjusted, all participants | Power-law Mixed | -0.093 [-0.290–0.104] | 430.3 | 441.3 |
| A.Idaho | Unadjusted, all participants | Cubic Spline Mixed |  | 619.5 | 638.3 |
| A.Idaho | Unadjusted, all participants | Linear Mixed | 0.009 [-0.011–0.028] | 634.4 | 647.0 |
| A.Idaho | Unadjusted, all participants | Power-law Mixed | -0.156 [-0.387–0.075] | 463.8 | 474.8 |
| A.Kansas | Unadjusted, all participants | Cubic Spline Mixed |  | 571.8 | 590.6 |
| A.Kansas | Unadjusted, all participants | Linear Mixed | 0.005 [-0.011–0.022] | 575.3 | 587.9 |
| A.Kansas | Unadjusted, all participants | Power-law Mixed | -0.032 [-0.232–0.168] | 422.0 | 433.0 |
| A.Michigan | Unadjusted, all participants | Cubic Spline Mixed |  | 558.4 | 577.2 |
| A.Michigan | Unadjusted, all participants | Linear Mixed | 0.005 [-0.011–0.021] | 578.1 | 590.7 |
| A.Michigan | Unadjusted, all participants | Power-law Mixed | -0.182 [-0.370–0.006] | 426.7 | 437.6 |
| A.Singapore | Unadjusted, all participants | Cubic Spline Mixed |  | 573.7 | 592.5 |
| A.Singapore | Unadjusted, all participants | Linear Mixed | 0.016 [-0.000–0.033] | 585.0 | 597.5 |
| A.Singapore | Unadjusted, all participants | Power-law Mixed | -0.005 [-0.200–0.191] | 432.9 | 443.8 |
| B.Colorado | Unadjusted, all participants | Cubic Spline Mixed |  | 474.6 | 493.4 |
| B.Colorado | Unadjusted, all participants | Linear Mixed | 0.006 [-0.007–0.019] | 479.8 | 492.4 |
| B.Colorado | Unadjusted, all participants | Power-law Mixed | -0.060 [-0.199–0.080] | 349.2 | 360.1 |
| B.Phuket | Unadjusted, all participants | Cubic Spline Mixed |  | 518.4 | 537.2 |
| B.Phuket | Unadjusted, all participants | Linear Mixed | 0.010 [-0.004–0.024] | 521.5 | 534.0 |
| B.Phuket | Unadjusted, all participants | Power-law Mixed | 0.033 [-0.144–0.209] | 392.5 | 403.5 |
| A.Brisbane | Adjusted, all participants | Cubic Spline Mixed |  | 548.1 | 573.2 |
| A.Brisbane | Adjusted, all participants | Linear Mixed | 0.013 [-0.004–0.029] | 556.0 | 574.8 |
| A.Brisbane | Adjusted, all participants | Power-law Mixed | -0.082 [-0.275–0.111] | 417.5 | 433.9 |
| A.Idaho | Adjusted, all participants | Cubic Spline Mixed |  | 600.5 | 625.6 |
| A.Idaho | Adjusted, all participants | Linear Mixed | 0.010 [-0.009–0.029] | 615.3 | 634.1 |
| A.Idaho | Adjusted, all participants | Power-law Mixed | -0.148 [-0.374–0.078] | 450.9 | 467.3 |
| A.Kansas | Adjusted, all participants | Cubic Spline Mixed |  | 562.4 | 587.5 |
| A.Kansas | Adjusted, all participants | Linear Mixed | 0.006 [-0.010–0.022] | 566.5 | 585.4 |
| A.Kansas | Adjusted, all participants | Power-law Mixed | -0.052 [-0.250–0.146] | 419.6 | 436.0 |
| A.Michigan | Adjusted, all participants | Cubic Spline Mixed |  | 528.4 | 553.5 |
| A.Michigan | Adjusted, all participants | Linear Mixed | 0.006 [-0.010–0.022] | 547.8 | 566.6 |
| A.Michigan | Adjusted, all participants | Power-law Mixed | -0.175 [-0.359–0.010] | 402.7 | 419.1 |
| A.Singapore | Adjusted, all participants | Cubic Spline Mixed |  | 539.0 | 564.1 |
| A.Singapore | Adjusted, all participants | Linear Mixed | 0.016 [-0.001–0.032] | 551.3 | 570.1 |
| A.Singapore | Adjusted, all participants | Power-law Mixed | -0.045 [-0.240–0.150] | 406.8 | 423.2 |
| B.Colorado | Adjusted, all participants | Cubic Spline Mixed |  | 427.5 | 452.5 |
| B.Colorado | Adjusted, all participants | Linear Mixed | 0.004 [-0.009–0.017] | 433.1 | 452.0 |
| B.Colorado | Adjusted, all participants | Power-law Mixed | -0.082 [-0.220–0.055] | 322.4 | 338.8 |
| B.Phuket | Adjusted, all participants | Cubic Spline Mixed |  | 494.7 | 519.8 |
| B.Phuket | Adjusted, all participants | Linear Mixed | 0.011 [-0.003–0.025] | 497.8 | 516.6 |
| B.Phuket | Adjusted, all participants | Power-law Mixed | 0.028 [-0.144–0.200] | 375.8 | 392.2 |
| A.Brisbane | Adjusted, baseline titer at least 1:40 | Cubic Spline Mixed |  | 401.6 | 424.9 |
| A.Brisbane | Adjusted, baseline titer at least 1:40 | Linear Mixed | 0.012 [-0.006–0.029] | 407.5 | 425.0 |
| A.Brisbane | Adjusted, baseline titer at least 1:40 | Power-law Mixed | -0.095 [-0.301–0.112] | 308.0 | 323.2 |
| A.Idaho | Adjusted, baseline titer at least 1:40 | Cubic Spline Mixed |  | 449.4 | 472.9 |
| A.Idaho | Adjusted, baseline titer at least 1:40 | Linear Mixed | 0.003 [-0.017–0.023] | 461.1 | 478.7 |
| A.Idaho | Adjusted, baseline titer at least 1:40 | Power-law Mixed | -0.232 [-0.467–0.004] | 338.1 | 353.4 |
| A.Kansas | Adjusted, baseline titer at least 1:40 | Cubic Spline Mixed |  | 438.0 | 461.8 |
| A.Kansas | Adjusted, baseline titer at least 1:40 | Linear Mixed | 0.002 [-0.016–0.019] | 441.7 | 459.5 |
| A.Kansas | Adjusted, baseline titer at least 1:40 | Power-law Mixed | -0.021 [-0.226–0.184] | 339.3 | 354.9 |
| A.Michigan | Adjusted, baseline titer at least 1:40 | Cubic Spline Mixed |  | 406.0 | 429.8 |
| A.Michigan | Adjusted, baseline titer at least 1:40 | Linear Mixed | 0.002 [-0.014–0.019] | 422.2 | 440.0 |
| A.Michigan | Adjusted, baseline titer at least 1:40 | Power-law Mixed | -0.216 [-0.402–-0.029] | 312.9 | 328.4 |
| A.Singapore | Adjusted, baseline titer at least 1:40 | Cubic Spline Mixed |  | 292.8 | 315.3 |
| A.Singapore | Adjusted, baseline titer at least 1:40 | Linear Mixed | 0.013 [-0.002–0.028] | 299.0 | 315.9 |
| A.Singapore | Adjusted, baseline titer at least 1:40 | Power-law Mixed | -0.033 [-0.182–0.115] | 230.3 | 244.9 |
| B.Colorado | Adjusted, baseline titer at least 1:40 | Cubic Spline Mixed |  | 226.2 | 248.1 |
| B.Colorado | Adjusted, baseline titer at least 1:40 | Linear Mixed | 0.012 [-0.000–0.024] | 230.4 | 246.8 |
| B.Colorado | Adjusted, baseline titer at least 1:40 | Power-law Mixed | -0.003 [-0.155–0.149] | 185.9 | 199.9 |
| B.Phuket | Adjusted, baseline titer at least 1:40 | Cubic Spline Mixed |  | 330.3 | 353.0 |
| B.Phuket | Adjusted, baseline titer at least 1:40 | Linear Mixed | 0.011 [-0.006–0.027] | 332.6 | 349.6 |
| B.Phuket | Adjusted, baseline titer at least 1:40 | Power-law Mixed | 0.068 [-0.124–0.260] | 256.9 | 271.5 |

Table S6: Multivariable antibody waning MN model results for HIVE among participant with baseline titer >=1:40

| Strain | Model | Change by Month (95% CI) | AIC | BIC |
| --- | --- | --- | --- | --- |
| A/H1N1/Michigan | Cubic Spline Mixed |  | 519.6 | 544.6 |
| A/H1N1/Michigan | Linear Mixed | 0.005 [0.004–0.007] | 538.2 | 556.9 |
| A/H1N1/Michigan | Power-law Mixed | -0.216 [-0.402–-0.029] | 397.1 | 413.4 |
| A/H3N2/Singapore | Cubic Spline Mixed |  | 528.4 | 553.4 |
| A/H3N2/Singapore | Linear Mixed | 0.006 [0.005–0.008] | 540.9 | 559.6 |
| A/H3N2/Singapore | Power-law Mixed | -0.033 [-0.182–0.115] | 398.5 | 414.9 |
| A/H3N2/Kansas | Cubic Spline Mixed |  | 549.6 | 574.6 |
| A/H3N2/Kansas | Linear Mixed | 0.002 [0.002–0.003] | 553.6 | 572.4 |
| A/H3N2/Kansas | Power-law Mixed | -0.021 [-0.226–0.184] | 409.9 | 426.3 |
| A/H1N1/Idaho | Cubic Spline Mixed |  | 591.8 | 616.8 |
| A/H1N1/Idaho | Linear Mixed | 0.003 [0.002–0.004] | 606.1 | 624.8 |
| A/H1N1/Idaho | Power-law Mixed | -0.232 [-0.467–0.004] | 445.5 | 461.9 |

Table S7: Antibody waning HAI model results for EPI-HK

| Strain | Type | Model | Change by month (95% CI) | AIC | BIC |
| --- | --- | --- | --- | --- | --- |
| A.Brisbane | Unadjusted, all participants | Cubic Spline Mixed |  | 647.7 | 669.0 |
| A.Brisbane | Unadjusted, all participants | Linear Mixed | -0.004 [-0.013–0.005] | 649.8 | 664.0 |
| A.Brisbane | Unadjusted, all participants | Power-law Mixed | 0.070 [-0.005–0.145] | 524.9 | 538.0 |
| A.Cambodia | Unadjusted, all participants | Cubic Spline Mixed |  | 563.3 | 584.7 |
| A.Cambodia | Unadjusted, all participants | Linear Mixed | -0.003 [-0.011–0.006] | 579.3 | 593.5 |
| A.Cambodia | Unadjusted, all participants | Power-law Mixed | -0.010 [-0.067–0.048] | 408.3 | 421.4 |
| A.Hawaii | Unadjusted, all participants | Cubic Spline Mixed |  | 470.1 | 491.5 |
| A.Hawaii | Unadjusted, all participants | Linear Mixed | 0.000 [-0.007–0.007] | 477.4 | 491.6 |
| A.Hawaii | Unadjusted, all participants | Power-law Mixed | 0.047 [-0.003–0.097] | 346.6 | 359.8 |
| A.Hong.Kong | Unadjusted, all participants | Cubic Spline Mixed |  | 785.3 | 806.7 |
| A.Hong.Kong | Unadjusted, all participants | Linear Mixed | -0.014 [-0.027–-0.002] | 789.5 | 803.7 |
| A.Hong.Kong | Unadjusted, all participants | Power-law Mixed | -0.067 [-0.174–0.039] | 634.6 | 647.7 |
| A.Kansas | Unadjusted, all participants | Cubic Spline Mixed |  | 365.4 | 386.8 |
| A.Kansas | Unadjusted, all participants | Linear Mixed | 0.001 [-0.005–0.006] | 364.2 | 378.4 |
| A.Kansas | Unadjusted, all participants | Power-law Mixed | 0.026 [-0.019–0.071] | 275.9 | 289.1 |
| A.Michigan | Unadjusted, all participants | Cubic Spline Mixed |  | 575.5 | 596.8 |
| A.Michigan | Unadjusted, all participants | Linear Mixed | -0.008 [-0.016–-0.001] | 577.4 | 591.6 |
| A.Michigan | Unadjusted, all participants | Power-law Mixed | -0.017 [-0.069–0.035] | 431.7 | 444.9 |
| A.Singapore | Unadjusted, all participants | Cubic Spline Mixed |  | 477.2 | 498.5 |
| A.Singapore | Unadjusted, all participants | Linear Mixed | -0.006 [-0.013–0.000] | 495.5 | 509.7 |
| A.Singapore | Unadjusted, all participants | Power-law Mixed | 0.008 [-0.047–0.063] | 397.6 | 410.7 |
| A.Victoria | Unadjusted, all participants | Cubic Spline Mixed |  | 566.5 | 587.9 |
| A.Victoria | Unadjusted, all participants | Linear Mixed | -0.008 [-0.017–0.001] | 567.2 | 581.4 |
| A.Victoria | Unadjusted, all participants | Power-law Mixed | -0.031 [-0.098–0.036] | 428.9 | 442.1 |
| B.Phuket | Unadjusted, all participants | Cubic Spline Mixed |  | 931.9 | 953.2 |
| B.Phuket | Unadjusted, all participants | Linear Mixed | -0.016 [-0.034–0.002] | 940.2 | 954.4 |
| B.Phuket | Unadjusted, all participants | Power-law Mixed | 0.046 [-0.088–0.180] | 704.7 | 717.9 |
| B.Washington | Unadjusted, all participants | Cubic Spline Mixed |  | 629.8 | 651.1 |
| B.Washington | Unadjusted, all participants | Linear Mixed | -0.005 [-0.014–0.004] | 630.5 | 644.7 |
| B.Washington | Unadjusted, all participants | Power-law Mixed | 0.016 [-0.056–0.088] | 486.8 | 499.9 |
| A.Brisbane | Adjusted, all participants | Cubic Spline Mixed |  | 3876.5 | 3923.4 |
| A.Brisbane | Adjusted, all participants | Linear Mixed | -0.004 [-0.006–-0.001] | 3898.1 | 3933.3 |
| A.Brisbane | Adjusted, all participants | Power-law Mixed | 0.071 [0.051–0.091] | 2789.8 | 2823.4 |
| A.Cambodia | Adjusted, all participants | Cubic Spline Mixed |  | 3332.0 | 3378.8 |
| A.Cambodia | Adjusted, all participants | Linear Mixed | -0.003 [-0.005–-0.001] | 3529.4 | 3564.5 |
| A.Cambodia | Adjusted, all participants | Power-law Mixed | -0.010 [-0.026–0.005] | 1724.9 | 1758.4 |
| A.Hawaii | Adjusted, all participants | Cubic Spline Mixed |  | 2421.7 | 2468.5 |
| A.Hawaii | Adjusted, all participants | Linear Mixed | -0.000 [-0.002–0.002] | 2519.2 | 2554.4 |
| A.Hawaii | Adjusted, all participants | Power-law Mixed | 0.047 [0.034–0.061] | 1196.3 | 1229.9 |
| A.Hong.Kong | Adjusted, all participants | Cubic Spline Mixed |  | 5489.1 | 5536.0 |
| A.Hong.Kong | Adjusted, all participants | Linear Mixed | -0.015 [-0.018–-0.011] | 5519.8 | 5554.9 |
| A.Hong.Kong | Adjusted, all participants | Power-law Mixed | -0.075 [-0.104–-0.046] | 4171.8 | 4205.3 |
| A.Kansas | Adjusted, all participants | Cubic Spline Mixed |  | 1500.6 | 1547.5 |
| A.Kansas | Adjusted, all participants | Linear Mixed | 0.000 [-0.001–0.002] | 1500.1 | 1535.3 |
| A.Kansas | Adjusted, all participants | Power-law Mixed | 0.021 [0.009–0.033] | 737.8 | 771.3 |
| A.Michigan | Adjusted, all participants | Cubic Spline Mixed |  | 2869.8 | 2916.7 |
| A.Michigan | Adjusted, all participants | Linear Mixed | -0.008 [-0.010–-0.006] | 2897.9 | 2933.0 |
| A.Michigan | Adjusted, all participants | Power-law Mixed | -0.015 [-0.029–-0.001] | 1422.7 | 1456.3 |
| A.Singapore | Adjusted, all participants | Cubic Spline Mixed |  | 2148.4 | 2195.3 |
| A.Singapore | Adjusted, all participants | Linear Mixed | -0.007 [-0.009–-0.005] | 2387.6 | 2422.8 |
| A.Singapore | Adjusted, all participants | Power-law Mixed | 0.006 [-0.009–0.021] | 1555.8 | 1589.3 |
| A.Victoria | Adjusted, all participants | Cubic Spline Mixed |  | 3637.6 | 3684.4 |
| A.Victoria | Adjusted, all participants | Linear Mixed | -0.009 [-0.011–-0.006] | 3641.1 | 3676.2 |
| A.Victoria | Adjusted, all participants | Power-law Mixed | -0.029 [-0.047–-0.011] | 2317.4 | 2351.0 |
| B.Phuket | Adjusted, all participants | Cubic Spline Mixed |  | 7361.5 | 7408.3 |
| B.Phuket | Adjusted, all participants | Linear Mixed | -0.017 [-0.022–-0.012] | 7424.3 | 7459.4 |
| B.Phuket | Adjusted, all participants | Power-law Mixed | 0.046 [0.010–0.082] | 5065.2 | 5098.7 |
| B.Washington | Adjusted, all participants | Cubic Spline Mixed |  | 3992.2 | 4039.1 |
| B.Washington | Adjusted, all participants | Linear Mixed | -0.005 [-0.008–-0.003] | 3994.2 | 4029.3 |
| B.Washington | Adjusted, all participants | Power-law Mixed | 0.016 [-0.003–0.036] | 2652.2 | 2685.7 |
| A.Brisbane | Adjusted, baseline titer at least 1:40 | Cubic Spline Mixed |  | 249.2 | 268.1 |
| A.Brisbane | Adjusted, baseline titer at least 1:40 | Linear Mixed | -0.010 [-0.037–0.017] | 254.8 | 269.1 |
| A.Brisbane | Adjusted, baseline titer at least 1:40 | Power-law Mixed | 0.157 [-0.035–0.349] | 204.0 | 216.7 |
| A.Cambodia | Adjusted, baseline titer at least 1:40 | Cubic Spline Mixed |  | 78.6 | 87.3 |
| A.Cambodia | Adjusted, baseline titer at least 1:40 | Linear Mixed | -0.066 [-0.108–-0.024] | 84.8 | 91.3 |
| A.Cambodia | Adjusted, baseline titer at least 1:40 | Power-law Mixed | -0.230 [-0.584–0.124] | 70.0 | 75.0 |
| A.Hawaii | Adjusted, baseline titer at least 1:40 | Cubic Spline Mixed |  | 81.9 | 91.3 |
| A.Hawaii | Adjusted, baseline titer at least 1:40 | Linear Mixed | -0.016 [-0.060–0.028] | 91.3 | 98.3 |
| A.Hawaii | Adjusted, baseline titer at least 1:40 | Power-law Mixed | 0.003 [-0.282–0.288] | 68.4 | 74.1 |
| A.Hong.Kong | Adjusted, baseline titer at least 1:40 | Cubic Spline Mixed |  | 262.1 | 282.0 |
| A.Hong.Kong | Adjusted, baseline titer at least 1:40 | Linear Mixed | -0.040 [-0.060–-0.019] | 269.9 | 284.9 |
| A.Hong.Kong | Adjusted, baseline titer at least 1:40 | Power-law Mixed | -0.060 [-0.258–0.138] | 220.0 | 233.3 |
| A.Kansas | Adjusted, baseline titer at least 1:40 | Cubic Spline Mixed |  | NA | NA |
| A.Kansas | Adjusted, baseline titer at least 1:40 | Linear Mixed | NA | NA | NA |
| A.Kansas | Adjusted, baseline titer at least 1:40 | Power-law Mixed | NA | NA | NA |
| A.Michigan | Adjusted, baseline titer at least 1:40 | Cubic Spline Mixed |  | 191.3 | 209.6 |
| A.Michigan | Adjusted, baseline titer at least 1:40 | Linear Mixed | -0.030 [-0.051–-0.010] | 198.1 | 211.8 |
| A.Michigan | Adjusted, baseline titer at least 1:40 | Power-law Mixed | -0.084 [-0.207–0.038] | 146.1 | 158.4 |
| A.Singapore | Adjusted, baseline titer at least 1:40 | Cubic Spline Mixed |  | 71.7 | 82.6 |
| A.Singapore | Adjusted, baseline titer at least 1:40 | Linear Mixed | -0.042 [-0.066–-0.019] | 78.1 | 86.3 |
| A.Singapore | Adjusted, baseline titer at least 1:40 | Power-law Mixed | -0.192 [-0.397–0.013] | 65.5 | 72.0 |
| A.Victoria | Adjusted, baseline titer at least 1:40 | Cubic Spline Mixed |  | 69.0 | 77.7 |
| A.Victoria | Adjusted, baseline titer at least 1:40 | Linear Mixed | -0.022 [-0.054–0.009] | 74.2 | 80.7 |
| A.Victoria | Adjusted, baseline titer at least 1:40 | Power-law Mixed | -0.063 [-0.348–0.223] | 59.8 | 64.8 |
| B.Phuket | Adjusted, baseline titer at least 1:40 | Cubic Spline Mixed |  | 541.9 | 566.1 |
| B.Phuket | Adjusted, baseline titer at least 1:40 | Linear Mixed | -0.029 [-0.053–-0.005] | 556.6 | 574.7 |
| B.Phuket | Adjusted, baseline titer at least 1:40 | Power-law Mixed | 0.068 [-0.087–0.224] | 411.9 | 428.5 |
| B.Washington | Adjusted, baseline titer at least 1:40 | Cubic Spline Mixed |  | 102.0 | 111.4 |
| B.Washington | Adjusted, baseline titer at least 1:40 | Linear Mixed | -0.064 [-0.120–-0.008] | 109.3 | 116.4 |
| B.Washington | Adjusted, baseline titer at least 1:40 | Power-law Mixed | -0.155 [-0.659–0.349] | 90.7 | 96.3 |

Table S8: Multivariable antibody waning MN model results for EPI-HK among participant with baseline titer >=1:40

| Strain | Model | Change by Month (95% CI) | AIC | BIC |
| --- | --- | --- | --- | --- |
| A/H1N1/Michigan | Cubic Spline Mixed |  | 152.6 | 166.1 |
| A/H1N1/Michigan | Linear Mixed | -0.018 [-0.033–-0.004] | 153.0 | 162.1 |
| A/H1N1/Michigan | Power-law Mixed | -0.143 [-0.325–0.039] | 155.3 | 164.4 |
| A/H3N2/Singapore | Cubic Spline Mixed |  | 173.8 | 187.4 |
| A/H3N2/Singapore | Linear Mixed | -0.026 [-0.040–-0.012] | 176.6 | 185.7 |
| A/H3N2/Singapore | Power-law Mixed | -0.060 [-0.194–0.073] | 173.9 | 182.9 |
| A/H3N2/Kansas | Cubic Spline Mixed |  | 180.8 | 194.4 |
| A/H3N2/Kansas | Linear Mixed | -0.004 [-0.024–0.017] | 182.4 | 191.5 |
| A/H3N2/Kansas | Power-law Mixed | -0.052 [-0.272–0.167] | 182.1 | 191.2 |
